# Ethnic disparities in immunisation: analyses of zero-dose prevalence in 64 low- and middle-income countries

**DOI:** 10.1101/2022.02.09.22270671

**Authors:** Bianca O Cata-Preta, Thiago M Santos, Andrea Wendt, Daniel R Hogan, Tewodaj Mengistu, Aluísio JD Barros, Cesar G Victora

## Abstract

**Background:** The Sustainable Development Goals (SDGs) recommend stratification of health indicators by ethnic group, yet there are few studies that have assessed if there are ethnic disparities in childhood immunisation in low- and middle-income countries (LMICs).

**Methods:** We identified 64 LMICs with standardized national surveys carried out since 2010, which provided information on ethnicity or a proxy variable and on vaccine coverage; 339 ethnic groups across the 64 countries were identified after excluding those with fewer than 50 children in the sample and countries with a single ethnic group. Lack of vaccination with diphtheria-pertussis-tetanus (DPT) vaccine – a proxy for no access to routine vaccination or ‘zero-dose’ status – was the outcome of interest. Differences among ethnic groups were assessed using a chi-squared test for heterogeneity. Additional analyses controlled for household wealth, maternal education and urban-rural residence.

**Findings:** The median gap between the highest and lowest zero-dose prevalence ethnic groups in all countries was equal to 10 percentage points (interquartile range 4-22; range 1 to 84) and the median ratio was 3.3 (interquartile range 1.8-6.7; range 1.1-30.4). In 35 of the 64 countries, there was significant heterogeneity in zero-dose prevalence among the ethnic groups. In most countries, adjustment for wealth, education and residence made little difference to the ethnic gaps, but in four countries (Angola, Benin, Nigeria, and Philippines) the high-low ethnic gap decreased by over 15 pp after adjustment. Children belonging to a majority group had 29% lower prevalence of zero-dose compared to the rest of the sample.

**Interpretation:** Statistically significant ethnic disparities in child immunisation were present in over half of the countries studied. Such inequalities have been seldom described in the published literature. Regular analyses of ethnic disparities are essential for monitoring trends, targeting resources and assessing the impact of health interventions to ensure zero-dose children are not left behind in the Sustainable Development Goals era.

**Funding:** Bill & Melinda Gates Foundation, Gavi, the Vaccine Alliance, Wellcome Trust, and Associação Brasileira de Saúde Coletiva.

## Backgorund

In 2020, there were 17 million children aged 12-23 months who failed to receive any doses of a DTP-containing vaccine. In alignment with the SDG motto of leaving no one behind, it is essential to identify these children through disaggregated analyses of existing datasets. Ethnicity within low- and middle-income countries (LMICs) is a likely determinant of access to services and immunization coverage. Yet, a PubMed search, searching articles from the last ten years, produced a single multicountry study investigating gaps in immunization coverage by ethnicity in 16 Latin American countries. We did not find any studies with data from multiple LMICs assessing differences in zero-dose prevalence by ethnicity.

### Added value of this study

We studied 64 LMICs and the median gap between the highest and lowest zero-dose prevalence ethnic group was 10 percent points. Gaps of 50 percent points or higher were found in five countries. In most countries, these differences persisted after adjustment for wealth, maternal education, and area of residence. We also found that children belonging to the majority ethnic group in a country tended to have lower zero-dose prevalence compared to the rest of the population.

### Implications of all the available evidence

In most LMICs, targeting by ethnic group is a potential strategy for reaching all children with immunizations. Our findings from 64 countries indicate which groups may be targeted. Regular analyses of ethnic disparities are also essential for monitoring trends over time and contribute to leaving no children behind with health interventions.

## Introduction

The 2030 Agenda for Sustainable Development, adopted by all United Nations Member States in 2015, has as its motto “leave no one behind” and thus prioritizes the elimination of within-country disparities due to income, gender, age, race, ethnicity, and other relevant characteristics.^1^ Specifically, Sustainable Development Goal (SDG) indicator 3.b.1 consists of the proportion of the target population covered by all vaccines included in their national program. Ensuring high and equitable coverage with vaccines is relevant to 14 of the 17 SDGs, and represents one of the “best buys” in global health.^2^ Supporting the SDGs, Immunization Agenda 2030 (IA 2030) envisions “a world where everyone, everywhere, at every age, fully benefits from vaccines to improve health and well-being”, and has as one of its targets the reduction of the number of zero-dose children globally by 50% by 2030.^3^

Analyses of inequalities in vaccine coverage within low- and middle-income countries (LMICs) are plentiful in the published literature, as well as in reports and websites from international organizations.^4^ Yet, studies of inequalities among ethnic groups tend to be restricted to single-country analyses^5-7^; we found a single multicountry study on this topic, which was restricted to 16 countries from Latin America and the Caribbean, using data collected from 2004 to 2015. ^8^ In most countries, this study revealed the presence of ethnic gaps in vaccine coverage. These are not unexpected given that ethnicity is a complex construct associated with health outcomes due to differences in health beliefs and behaviours.^9^ Ethnicity is also relevant to the dissemination of health information. Often, ethnic groups differ in terms of unequal access to socio-economic opportunities and to health services.

Consistent with the SDG motto of leaving no one behind and IA 2030 goals, countries and international organizations have focused on identifying “zero-dose” children, that is, those who have failed to receive any routine vaccinations. ^10-12^ To better operationalize the concept, identifying “zero-dose” has evolved to focus on no-DPT children, i.e., those without any doses of the diphtheria-pertussis-tetanus-containing vaccine, because the full “zero-dose” indicator is challenging to measure with routine administrative data in many countries.

In this study, we systematically identified publicly available household sample surveys from LMICs with information on DPT vaccinations and on ethnicity or a proxy variable such as language spoken at home. We examined ethnic gaps in zero-dose prevalence as measured by lack of DPT vaccination among children aged 12-23 months, and we assessed whether these gaps could be explained by differences among ethnic groups in terms of household wealth, maternal education, or area of residence. We also assessed whether children belonging to the majority ethnic group in each country were more likely to be vaccinated than the remaining children in the country.

## Methods

### Data sources

We reviewed nationally representative surveys since 2010 and identified 65 countries with available information on immunisations and ethnicity or a proxy variable. For countries with more than one survey, the most recent was selected. One country, Moldova, was excluded because no no-DPT children were present in the sample. Therefore, we analysed survey data from 64 countries, 33 with Demographic and Health Surveys (DHS) and 31 with Multiple Indicator Cluster Surveys (MICS). Further information on the surveys is available elsewhere: DHS (https://dhsprogram.com/what-we-do/survey-Types/dHs.cfm) and MICS (http://mics.unicef.org/). Both survey programs are highly comparable in terms of sampling and indicators.^13, 14^

### Study samples

Our sample included children aged 12-23 months. In Bosnia and Herzegovina (2011), and Peru (2019), we studied children aged 18-29 months because the measles vaccine is given after 12 months of age, in contrast to most countries where it is given at 9-12 months. Although measles vaccination is not included in our analyses, the use of these age ranges makes our indicator consistent with results from national survey reports and from recent publications on lack of vaccination.^10, 11^

### Immunisation indicator

The outcome variable – no-DPT prevalence – was defined as the proportion of children without any doses of a DPT-containing vaccine, including tetravalent and pentavalent vaccines. As discussed above, we use no DPT as a proxy for zero-dose children.^15, 16^ Information on immunisation status was extracted for children from two sources: vaccination cards or, when the child did not have a card or it was unavailable at the time of interview, the mother’s or caregiver’s report. We treated children with missing information on immunization as not vaccinated.

### Ethnicity indicators

Within each sampled household, women aged 15-49 years (DHS) or the head of the household (MICS), provided information on ethnicity or a proxy variable. These included self-reported ethnicity in 45 surveys, language spoken at home or by the household head in 17 surveys, skin colour and caste in one survey each (Cuba and India, respectively). In Latin America and the Caribbean, for consistency with earlier analyses, we grouped the ethnic variable into three categories as follows: reference (mostly individuals with European, or mixed European and indigenous ancestry), indigenous and afrodescendants.^17^ Our results below show the ethnic group labels according to each survey dataset; some labels include more than one denomination, as was the case for Chad. We recoded groups with fewer than 50 children into country-specific “other” categories; if the “other group” still included fewer than 50 children, these were excluded from all analyses. For simplicity, we refer to ethnicity to indicate either ethnic group, language, skin colour or caste.

A detailed listing of the groups in each country is available in Supplementary Table 1. In 38 countries (59% of all countries), it was possible to identify a majority ethnic group comprising half or more of the children in the sample (Supplementary Table 5).

### Statistical analyses

We calculated crude and adjusted no-DPT prevalence and their 95% confidence intervals (95% CI) by ethnic group as the marginal means from the prediction of a Poisson model with robust variance^18^ in each country. The presence of variability among ethnic groups in a country was assessed using a chi-squared test for heterogeneity. The adjusted model test whether wealth, education and residence explained the differences in no-DPT prevalence among ethnic groups. For both crude and adjusted prevalence, we calculated the coefficient of variation (CV) and highlighted ethnic groups with more precise estimates (CV < 15%) when presenting the results. We compared no-DPT prevalence between the highest and lowest prevalence ethnic groups in each country using differences and ratios. The statistical significance of the difference in no-DPT prevalence between the highest and lowest groups was assessed with a F-test. We described the distribution of prevalence differences and ratios using the median, interquartile range (IQR) and range.

Also, we pooled all countries and fitted a Poisson model with robust variance^18^ and fixed effect for countries to calculate crude and adjusted no-DPT prevalence ratios between the majority ethnic group (reference group) and all remaining children from the same country. Pooled results were weighted by the national population of children aged 12-23 months in 2016 (the median year of the surveys covered) obtained from the World Bank Population Estimates and Projections.^19^

The adjustment variables included maternal education, area of residence, and wealth quintiles. Maternal education was coded in three groups based on self-report: none (no formal education); primary (any primary education, including completed primary education); secondary or higher (any secondary education, including completed secondary education and partial or full higher education). Urban or rural residence was coded according to country-specific delimitations at the time of the survey. Household wealth indices included in the DHS and MICS datasets were used in the analyses. These were derived using principal component analyses of household assets and characteristics of the building, presence of electricity, water supply and sanitary facilities, among other variables associated with wealth. Because relevant assets may vary in urban and rural households, separate principal component analyses are carried out in each area, which are later combined into a single score using a scaling procedure to allow comparability between urban and rural households.^20^

The analyses were carried out with Stata (StataCorp. 2019. Stata Statistical Software: Release 17. College Station, TX: StataCorp LLC) and R (R Core Team, 2020, version 4.1.0. R Foundation for Statistical Computing, Vienna, Austria) and accounted for the multi-stage survey design and sampling weights.

### Ethical aspects

Ethical clearance was the responsibility of the institutions that administered the surveys and all analyses relied on anonymized databases.

## Results

Sixty-four countries with 339 ethnic groups and a total of 168,846 children were studied. The number of ethnic groups ranged from 20 in Uganda to 2 in Costa Rica, Cuba, Dominican Republic, Guatemala, Iraq, Kyrgyzstan, Mexico, Myanmar, North Macedonia, Paraguay, Peru, South Africa, Suriname, Tajikistan, Thailand, Timor-Leste, Turkmenistan and Vietnam (Table 1). In 38 countries, 50% or more of the children belonged to a single group, referred to as a majority group (Supplementary Table 5).

**Table 1.**
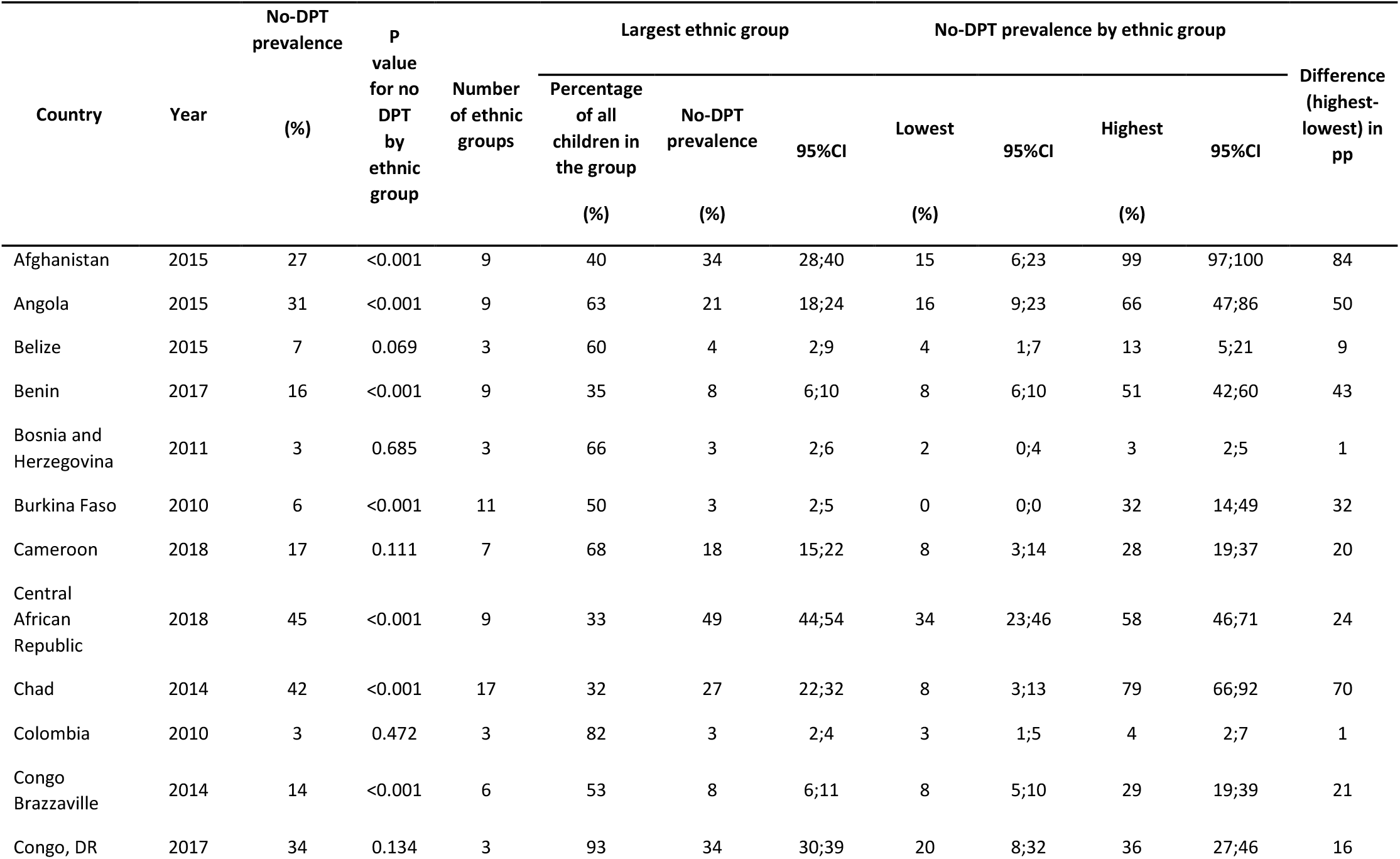
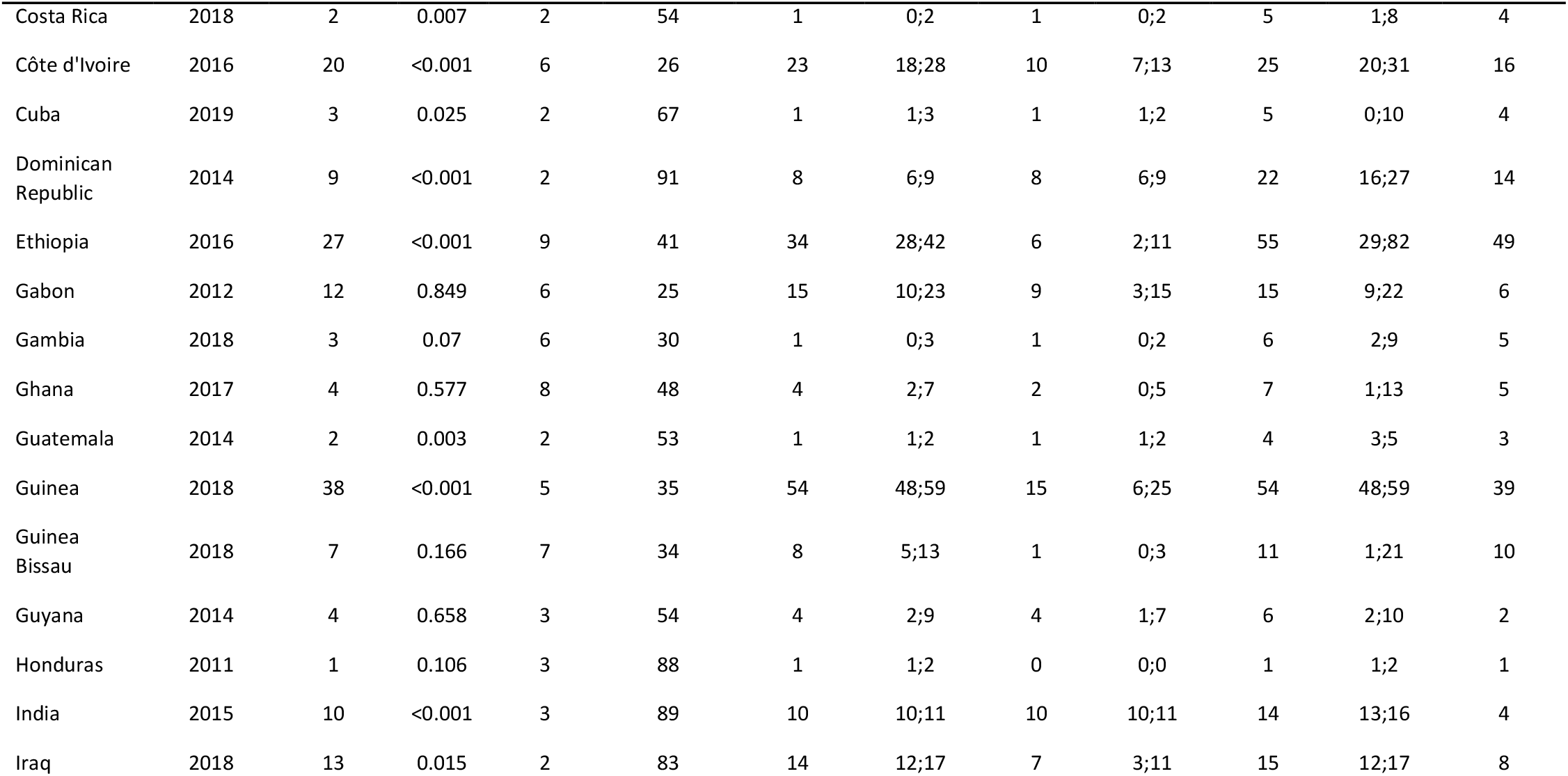
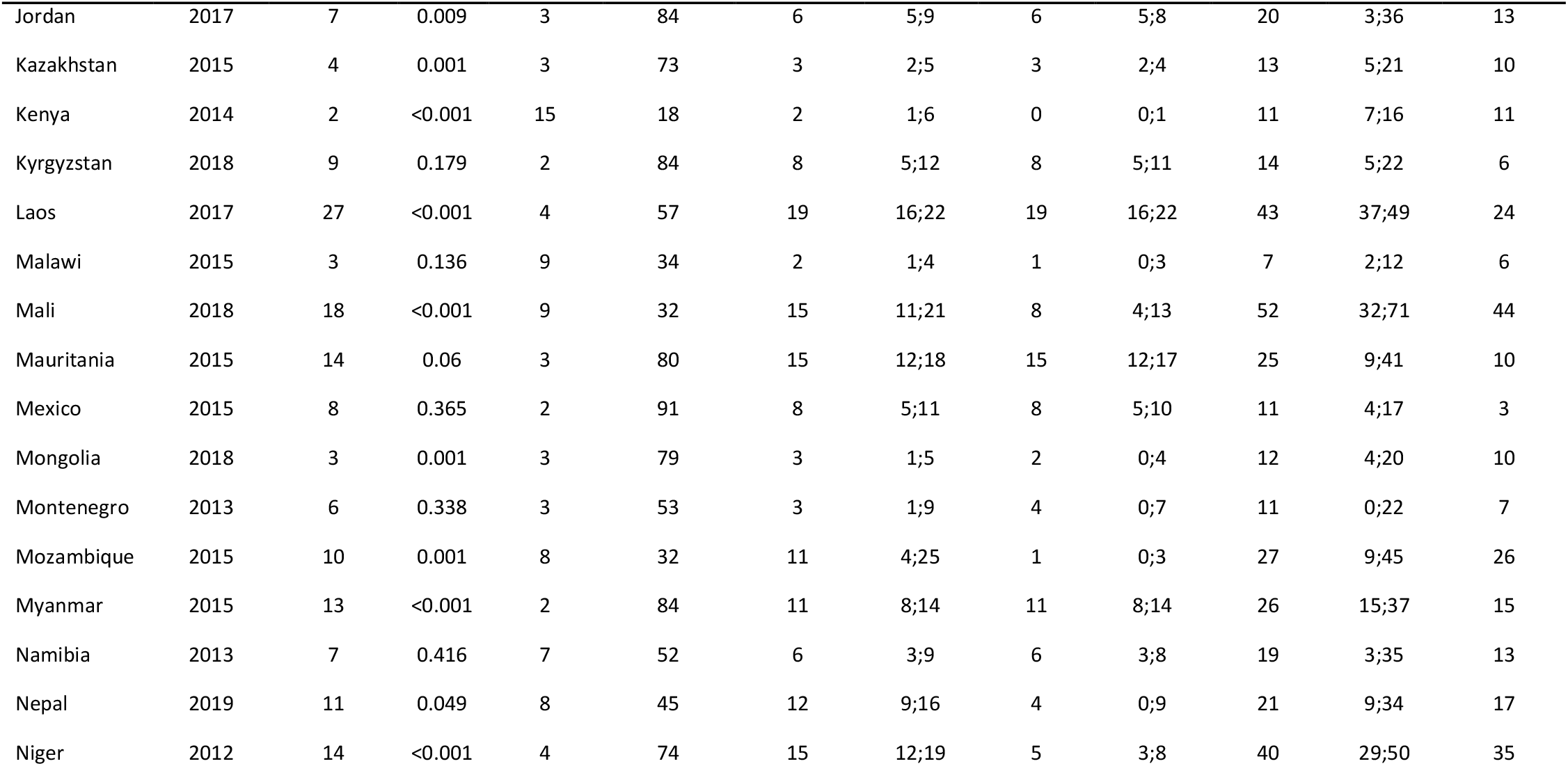
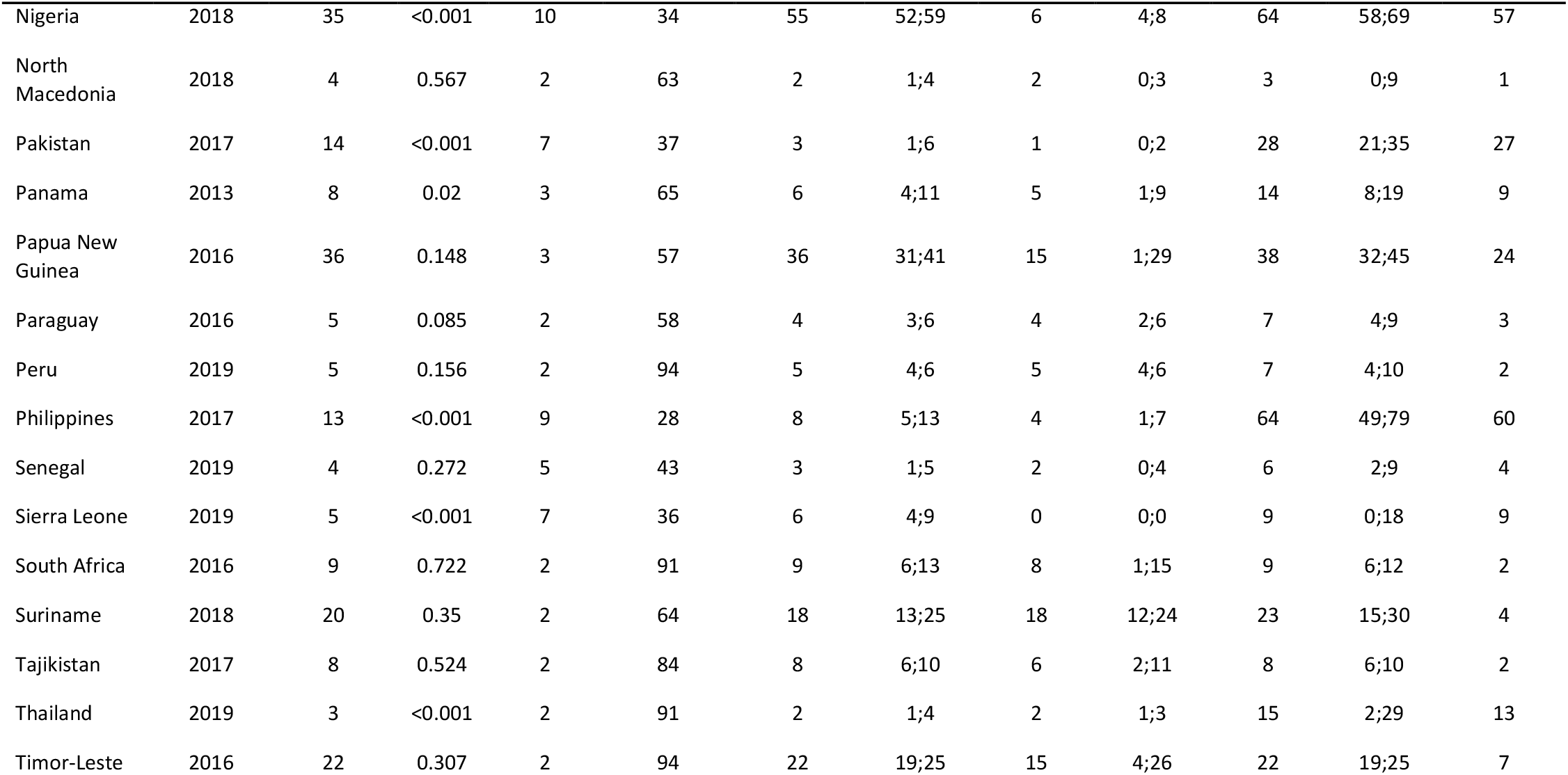
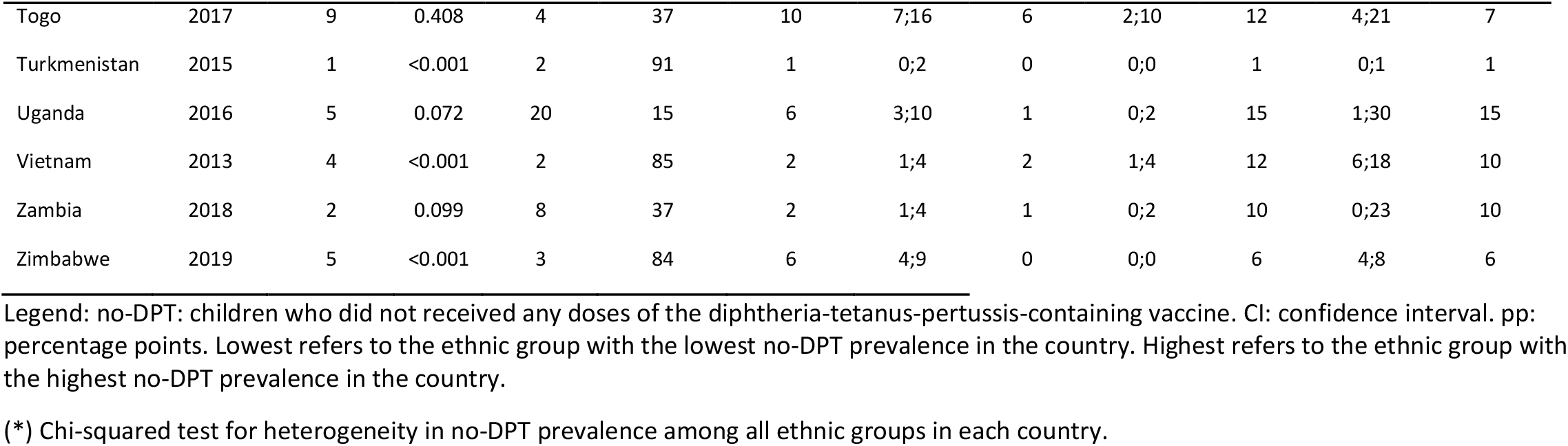
Countries included in the analyses, number of ethnic groups by country and no-DPT prevalence for ethnic groups. All values are percentages unless otherwise stated.

Table 1 presents no-DPT prevalence in each country, in the largest ethnic group and in the lowest and highest prevalence (in terms of no-DPT) ethnic groups, as well as differences between the extreme groups in percentage points (pp), and the p-value for the chi-test assessing heterogeneity in no-DPT prevalence among ethnic groups in each country. Supplementary Tables 2 and 3 show more detailed results, including the type of stratification variable (ethnicity, language, caste or skin colour), number of children, median no-DPT prevalence and high-low prevalence ratio for each country and no-DPT prevalence by ethnicity.

The median high-low no-DPT prevalence difference between ethnic groups was equal to 10 percentage points (interquartile range 4-22). Large prevalence gaps between extreme groups were observed in Afghanistan (83 pp), Chad (71 pp), Nigeria (57 pp), Philippines (60 pp), and Angola (50 pp) (Table 1). The median high-low prevalence ratio in all countries was 3.3 (interquartile range 1.8-6.7) (Supplementary Table 2). In 35 of the 64 countries, there was significant heterogeneity in no-DPT prevalence among ethnic groups (Table 1 and Supplementary Table 3). Among the remaining 29 countries, there were three (Central African Republic, Gambia and Papua New Guinea) where the 95% CIs of the two extreme groups did not overlap, although the overall test for heterogeneity showed p values above 0.05 (Supplementary Table 3). Among these countries, the difference between the extreme groups ranged from 5 pp in Gambia (between ethnic groups Mandinka and other) to 24 pp in Central African Republic (between ethnic groups Mandja and Yakoma/Sango) (Table 1 and Supplementary Table 3).

In 25 countries, at least one ethnic group presented no-DPT prevalence 10 pp or more above the national average (Table 1). Fifty-six ethnic groups in 15 countries showed 95% confidence intervals for no-DPT prevalence that were fully above 20% (Supplementary Table 3). At the other end of the prevalence range, 119 ethnic groups from 40 countries showed 95% confidence intervals below 10%.

Figure 1 shows no-DPT prevalence by ethnic groups in the 64 countries grouped by region of the world. Two lines are shown per country, the top one with the crude and another with the adjusted prevalence levels. The national prevalence is shown as a red circle. More precise prevalence estimates (CV <15%) are shown as black circles, and less precise estimates as grey circles. Full results and confidence intervals are shown in Supplementary Table 3, to which readers may refer to if interested in how adjustment influenced no-DPT prevalence in each country.

**Figure 1.**
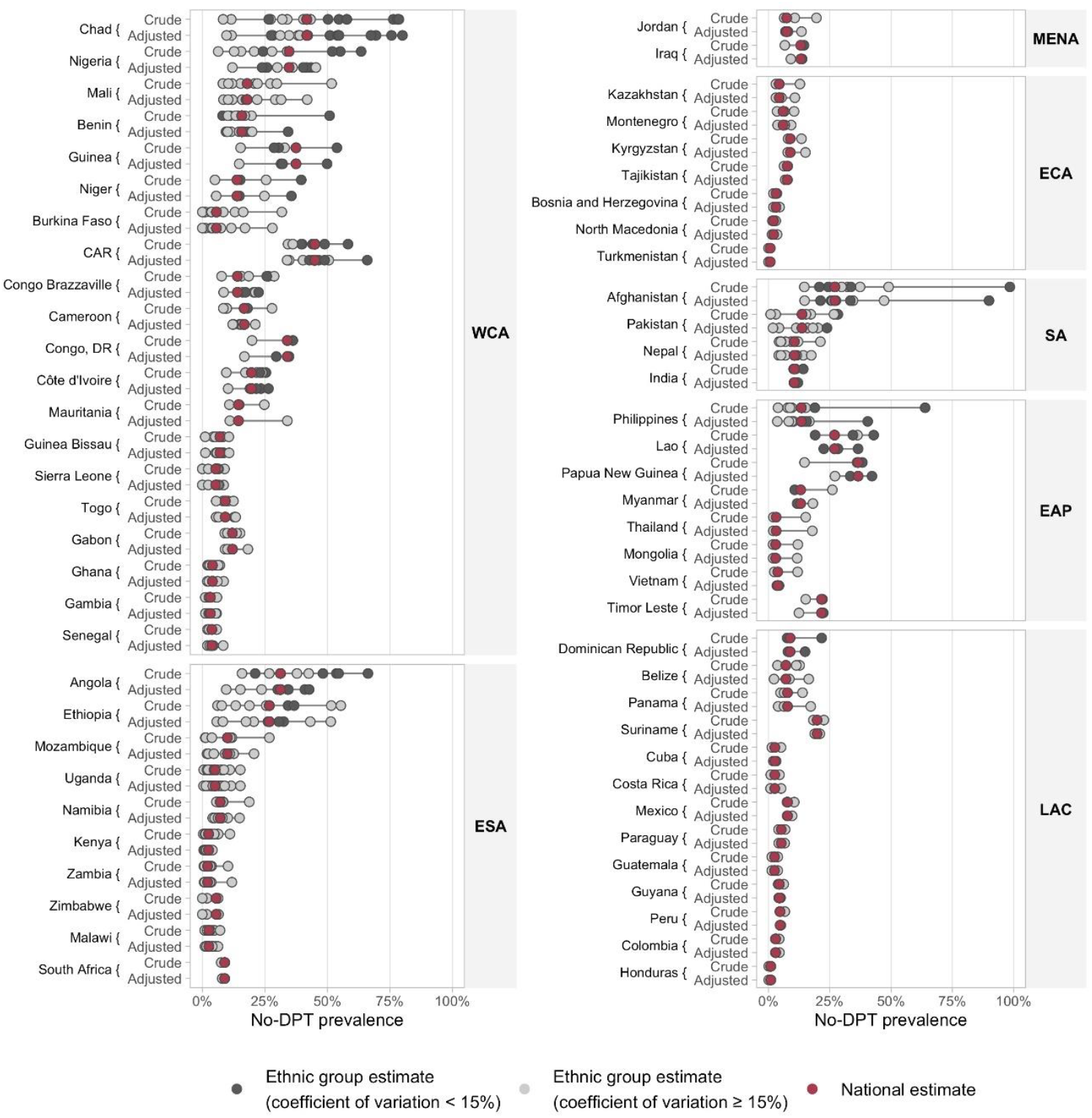
No-DPT prevalence by ethnic group in 64 countries. Black circles represent groups for which the coefficient of variation for prevalence was less than 15%. WCA: West and Central Africa. ESA: Eastern and South Africa. MENA: Middle East and North Africa. ECA: Europe and Central Asia. SA: South Asia. EAP: East Asia and The Pacific. LAC: Latin America and Caribbean. No-DPT: children who did not received any doses of the diphtheria-tetanus-pertussis-containing vaccine.

Adjustment for household wealth, maternal education and urban-rural residence made little difference in no-DPT prevalence gap in most countries. Among the 35 countries with statistically significant ethnic differences in the crude analyses, 26 remained significant in the adjusted analyses (Supplementary Table 3). Six countries with nonsignificant differences in the crude analysis presented p values under 0.05 in the adjusted analysis: Belize, Central African Republic, Mauritania, Papua New Guinea, Togo and Uganda.

In 19 countries the high-low ethnic gap changed by more than 5 pp (17 countries showing a reduction and two an increase in the gaps) after adjustment for covariates (Figure 1). The changes were greater than 15pp in Angola (17pp), Benin (18pp), Nigeria (26pp) and the Philippines (24pp), in all of which the gaps were narrowed down after adjustment. Increases in the ethnic gaps above 5 pp due to adjustment were only seen in the Central African Republic (7pp) and Mauritania (9pp).

We zoomed in on the 10 countries with the highest absolute gap in terms of no-DPT prevalence (Figure 2). Afghanistan and the Philippines stand out for each having a group (Nuristani and Maranao, respectively) with markedly higher no-DPT prevalence than any other ethnic group. Results for all countries are presented in Supplementary Figure 1 and Supplementary Table 3. Some ethnic groups are present in more than one country. For example, the Baloch or Baluchi show high no-DPT prevalence in both Afghanistan (49.0%) and Pakistan (27.9%). The Fula (also Peulh, Fulani, Fullah and similar denominations) are present in ten countries in the analyses, with no-DPT prevalence ranging widely from 0.0% in Sierra Leone to 63.5% in Nigeria. Supplementary table 4 shows these and other examples of groups with no-DPT prevalence higher than 10% present in more than one country.

**Figure 2.**
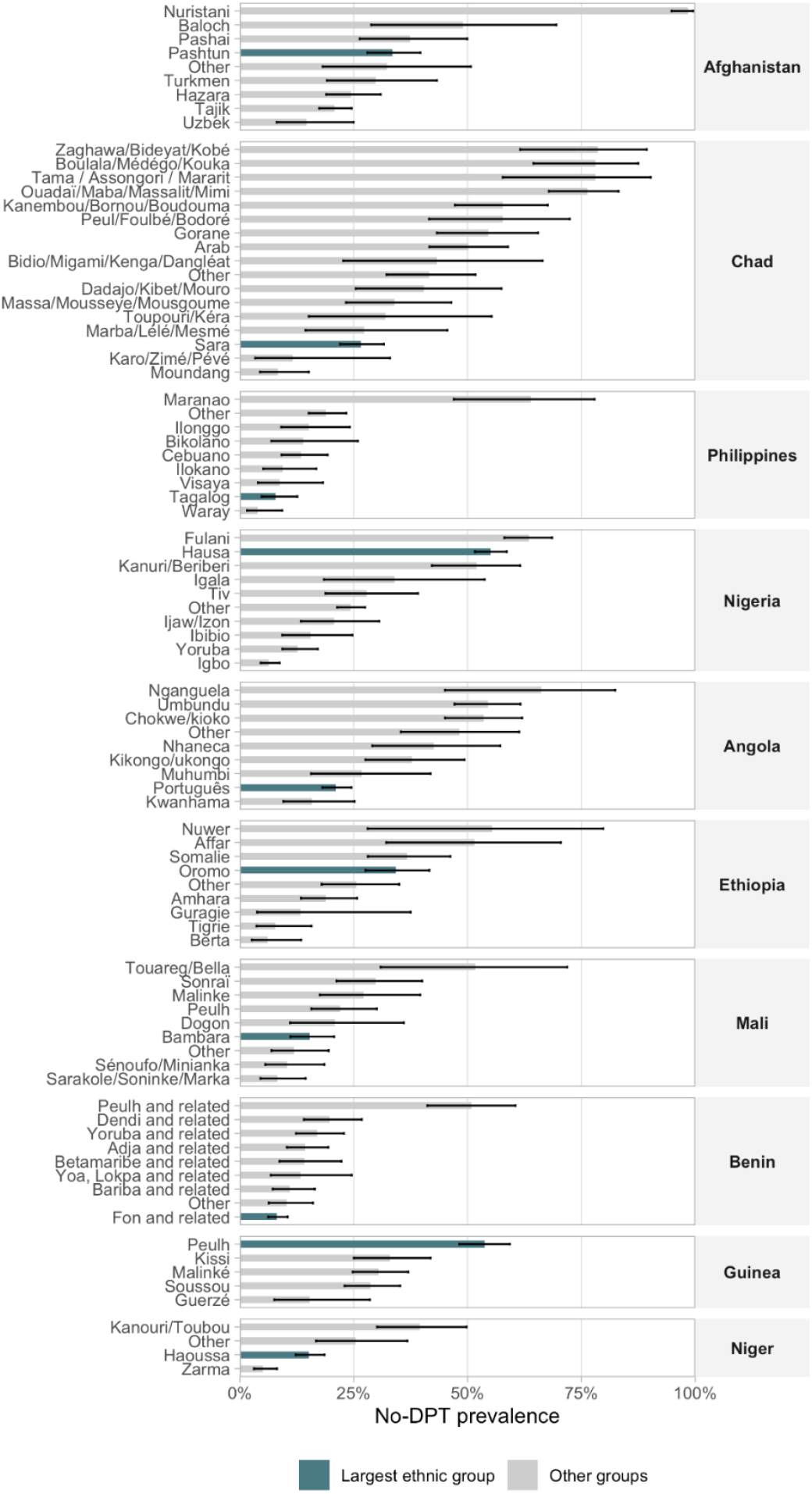
No-DPT prevalence according to ethnic group in the ten countries with the highest absolute ethnic gaps. No-DPT: children who did not received any doses of the diphtheria-tetanus-pertussis-containing vaccine.

Among the 40 Gavi eligible countries in the sample, 72.5% had significant ethnic gaps in no-DPT prevalence between the extreme groups, with a median gap of 15.6 pp in Gavi countries and 5.2 pp in non-Gavi eligible countries. The highest median differences were found in the South Asia (22.1 pp) and West and Central Africa (17.9 pp) regions and in countries with more than eight ethnic groups (43.5 pp). Supplementary table 6 shows the median no-DPT prevalence and the median high-low ethnic gaps according to Gavi eligibility status, Unicef world regions, World Bank country income groups, number of ethnic groups, and three ranges of no-DPT prevalence.

Lastly, we identified the 38 countries in which a single ethnic group comprised most of the children in the sample and compared their no-DPT prevalence with all other groups combined, using national child population weights for pooling results. Children from the majority ethnicity tended to have lower no-DPT prevalence than those from the remaining groups in the same country. The crude prevalence ratio in the majority group relative to the rest of the population was equal to 0.71 (95% CI 0.66; 0.77) while after adjustment for the three covariates the ratio changed to 0.82 (95% CI 0.76; 0.88).

## Discussion

We believe that this is the largest ever set of analyses on immunisation coverage according to ethnicity, covering 339 ethnic groups in 64 countries. Our results show important variations in no-DPT prevalence by ethnicity, with a median gap of 10 percentage points (and a median ratio of 3.3 times) between the highest- and lowest-prevalence groups in all countries studied. In 35 countries, there was statistically significant heterogeneity by ethnicity. Five countries showed gaps of 50 pp or more between their extreme groups. Our findings confirm the importance of subnational analyses with ethnic stratification. For example, in the Philippines the national no-DPT prevalence was 13% whereas the prevalence in ethnic groups ranged from 3.9% and 63.9%.

We also showed that the largest ethnic group in each sample was often not the lowest-prevalence group (Figure 2 and Supplementary Figure 1), but when all other groups were pooled the majority ethnicity tended to have higher DPT coverage than the remaining population as a group.

Studies from single low- and middle-income countries have reported ethnic differences in immunisation coverage, as was the case in studies from China, Kenya, the Philippines, and Pakistan.^5-7, 21^ We were only able to identify one multicountry study on immunisation coverage - in this case, with three DPT doses - by ethnic group; the analyses included 16 countries from Latin America and the Caribbean and relied on data collected from 2004 to 2015. ^8^ In three countries (Nicaragua, Panama, and Paraguay), indigenous children had significantly lower coverage than the reference group comprised of children of European or mixed ancestry. It should be noted, however, that immunisation coverage tends to be much higher in Latin America than in most LMICs.^22^ None of the studies identified in our literature search reported on no-DPT children according to ethnicity. Given the current emphasis on reaching zero-dose, there is a clear need for such studies to guide policy.

The literature, mostly from high-income countries, suggests that while adjusting for sociodemographic variables when comparing health outcomes among ethnic groups often attenuates disparities, these still persist.^23^ In our own analyses, ethnic gaps did not change markedly in most countries after adjustment for maternal education, household wealth and urban-rural residence. The exceptions included Angola, Benin, Nigeria and the Philippines, where the results suggest that socioeconomic factors account for a substantial proportion of the gaps. In many countries where the gaps persisted after adjustment, ethnic-based discrimination affecting the deployment and population access to essential services may account for much of the observed disparities. These differences could also reflect subnational variations in access, as some ethnic groups are highly concentrated in specific areas. For example, in Kenya, the largest no-DPT prevalence was found among Somali children who live in the Northeast of the country, and in the Philippines the Maranao children - who inhabit a well-delimited area of Mindanao island - show much higher no-DPT prevalence that in any other ethnic group in the country.

Our analyses have limitations, which include the use of self-reported ethnicity or proxy variables; this also applies to most studies of ethnic disparities in health.^24^ The way by which different ethnic groups were classified depended upon the agencies that developed questionnaires for each country, which may not have used consistent approaches, as is suggested by the wide variability in the number of groups among countries. Also, many survey datasets include some groups labelled as “other ethnicities”; due to sample size limitations, we also included in this category additional ethnic groups with fewer than 50 children in the sample. A particular case is that of India, where the ethnicity variable included only three groups: (any) caste, no caste or tribe, and tribe, with respectively 89.0%, 3.8% and 7.2% of all children. This classification showed that no-DPT prevalence range, from 10.2% among the former to 14.2% among the latter, but further breakdown showing the main castes would have been useful.

Our option for not reporting estimates for groups with small numbers of children has led to the omission of some potentially informative ethnic groups in some countries, for example whites in South Africa. In addition, there may be inconsistencies between successive surveys in some countries – for example the Nigeria 2016 MICS recognized only four groups, whereas the 2018 DHS used in the present analyses identified 10 groups plus an “other” category (Supplementary Table 1). One should also note that some ethnic groups – such as nomads or those living in conflict-afflicted areas – may be underrepresented in the sample. An additional limitation refers to the fact that surveys included in the analyses took place over a nine-year period, although we gave preference to more recent surveys when more than one existed for the same country. For countries without recent surveys, our findings may fail to describe the current situation.

In as much as we would like to calculate summary measures of inequality in order to rank countries according to the overall magnitude of ethnic gaps, such measures tend to show higher values in countries with many ethnic groups than for countries with few groups. In our analyses, significant differences between the highest and lowest ethnic groups in terms of zero-dose prevalence (p<0.05) were observed in 45% of countries with 2-3 groups, 61% of those with 4-8 groups, and in all but one countries with 9 or more groups (Supplementary Table 6). This limitation affects all summary measures of inequality for unordered categories.^25, 26^

Our analyses are limited to countries with recent surveys providing data both on ethnicity and DPT coverage. We examined surveys from over 100 countries to identify 64 that could be included in the present analyses. Whether or not our results may be generalized to other LMICs is debatable, but the fact that most countries showed significant ethnic gaps in no-DPT prevalence suggests that such inequalities may be present in countries that were not studied.

The purpose of our analyses was to present a broad picture of inequalities according to ethnic groups in access to immunisation based on recent national surveys. We showed important gaps in the over half of all countries under study. A detailed examination of the national contexts in which these inequalities exist is beyond the scope of the present analyses, but we hope that our results will motivate national researchers and other country actors to delve deeper into these disparities and their determinants. Further research may include an examination of the drivers of immunisation inequalities in different countries and comparisons between countries with contrasting patterns of ethnic group inequalities. Given that ethnicity appears to be a significant predictor of immunisation status in many LMICs, we advocate for greater attention to recording ethnicity in surveys and in health information systems, so as to allow monitoring, targeting of interventions and evaluating the equity impact of health services and programs.

Ideally, equity-oriented health programming and research on health inequalities should rely on multiple stratification variables to best characterize households and individuals. Although wealth and educational inequalities are useful for advocacy purposes and for monitoring time trends, they are often insufficient for targeting interventions at specific groups, as the poor and uneducated may be spread throughout a country. Geographical inequalities are better suited for targeting, but within a given province or district there may be important disparities, as is the case for large metropolitan areas. We show that ethnicity is an important dimension of immunisation inequalities in many LMICs and therefore assessments conducted to plan programming to reach zero-dose children and missed communities should routinely consider ethnicity to ensure no child is left behind with immunisation in the SDG era.

## Data Availability

All data produced in the present study are available upon reasonable request to the authors

## Data sharing statement

All the analyses were carried out using publicly available datasets that can be obtained directly from the DHS (dhsprogram.com) the MICS (mics.unicef.org) websites.

## Acknowledgements

This paper was made possible with funds from Bill & Melinda Gates Foundation (Grant Number: OPP1199234), Gavi, the Vaccine Alliance, Wellcome Trust (Grant Number: 101815/Z/13/Z), and Associação Brasileira de Saúde Coletiva.

## Contributors

All authors conceptualized the paper. BCP, TMS and AW conducted the analyses and verified the underlying data, with support from CGV and AJDB. All authors interpreted the results. BCP, AW and CGV prepared the first draft of the manuscript, which was revised and edited by all other authors. All authors read and approved the final manuscript.

## References

1. United Nations. Sustainable Development Goals. https://www.un.org/sustainabledevelopment/. Accessed Oct 27, 2021.

2. Gavi - The Vaccine Alliance. Sustainable Development Goals. https://www.gavi.org/our-alliance/global-health-development/sustainable-development-goals. Accessed 27 Oct 2021.

3. World Health Organization. Immunisation agenda 2030: a global strategy to leave no one behind. Geneva: WHO; 2020.

4. World Health Organization. State of inequality: childhood immunization. Geneva; 2016.

5. Bondy JN, Thind A, Koval JJ, Speechley KN. Identifying the determinants of childhood immunization in the Philippines. Vaccine. 2009;27(1):169–75. 10.1016/j.vaccine.2008.08.042.

6. Huang Y, Shallcross D, Pi L, Tian F, Pan J, Ronsmans C. Ethnicity and maternal and child health outcomes and service coverage in western China: a systematic review and meta-analysis. The Lancet Global health. 2018;6(1):e39–e56. 10.1016/s2214-109x(17)30445-x.

7. Siddiqui NT, Owais A, Agha A, Karim MS, Zaidi AK. Ethnic disparities in routine immunization coverage: a reason for persistent poliovirus circulation in Karachi, Pakistan? Asia-Pacific journal of public health. 2014;26(1):67–76. 10.1177/1010539513475648.

8. Mesenburg MA, Restrepo-Mendez MC, Amigo H, Balandran AD, Barbosa-Verdun MA, Caicedo-Velasquez B, et al. Ethnic group inequalities in coverage with reproductive, maternal and child health interventions: cross-sectional analyses of national surveys in 16 Latin American and Caribbean countries. Lancet Global Health. 2018;6(8):E902–E13. 10.1016/s2214-109x(18)30300-0.

9. Chaturvedi N. Ethnicity as an epidemiological determinant--crudely racist or crucially important? International journal of epidemiology. 2001;30(5):925–7. 10.1093/ije/30.5.925.

10. Cata-Preta BO, Santos TM, Mengistu T, Hogan DR, Barros AJD, Victora CG. Zero-dose children and the immunisation cascade: Understanding immunisation pathways in low and middle-income countries. Vaccine. 2021;39(32):4564–70. 10.1016/j.vaccine.2021.02.072.

11. Santos TM, Cata-Preta BO, Victora CG, Barros AJD. Finding Children with High Risk of Non-Vaccination in 92 Low- and Middle-Income Countries: A Decision Tree Approach. Vaccines. 2021;9(6). 10.3390/vaccines9060646.

12. Bosch-Capblanch X, Banerjee K, Burton A. Unvaccinated children in years of increasing coverage: how many and who are they? Evidence from 96 low- and middle-income countries. Trop Med Int Health. 2012;17(6):697–710. 10.1111/j.1365-3156.2012.02989.x.

13. Corsi DJ, Neuman M, Finlay JE, Subramanian SV. Demographic and health surveys: a profile. International journal of epidemiology. 2012;41(6):1602–13. 10.1093/ije/dys184.

14. Hancioglu A, Arnold F. Measuring coverage in MNCH: tracking progress in health for women and children using DHS and MICS household surveys. PLoS medicine. 2013;10(5):e1001391–e. 10.1371/journal.pmed.1001391.

15. Cherian T, Hwang A, Mantel C, Veira C, Malvolti S, MacDonald N, et al. Global Vaccine Action Plan lessons learned III: Monitoring and evaluation/accountability framework. Vaccine. 2020;38(33):5379–83. https://doi.org/10.1016/j.vaccine.2020.05.028.

16. Gavi - The Vaccine Alliance. The zero-dose child: explained. https://www.gavi.org/vaccineswork/zero-dose-child-explained. Accessed 27 Oct 2021.

17. Mesenburg MA, Restrepo-Mendez MC, Amigo H, Balandrán AD, Barbosa-Verdun MA, Caicedo-Velásquez B, et al. Ethnic group inequalities in coverage with reproductive, maternal and child health interventions: cross-sectional analyses of national surveys in 16 Latin American and Caribbean countries. The Lancet Global health. 2018;6(8):e902–e13. 10.1016/s2214-109x(18)30300-0.

18. Barros AJ, Hirakata VN. Alternatives for logistic regression in cross-sectional studies: an empirical comparison of models that directly estimate the prevalence ratio. BMC medical research methodology. 2003;3:21. 10.1186/1471-2288-3-21.

19. The World Bank. Health Nutrition and Population Statistics: Population estimates and projections. https://databank.worldbank.org/source/population-estimates-and-projections. Accessed October 10 2021.

20. Rutstein SO. The DHS Wealth Index: Approaches for rural and urban areas. 2008.

21. Masters NB, Wagner AL, Carlson BF, Muuo SW, Mutua MK, Boulton ML. Childhood vaccination in Kenya: socioeconomic determinants and disparities among the Somali ethnic community. International journal of public health. 2019;64(3):313–22. 10.1007/s00038-018-1187-2.

22. Restrepo-Mendez MC, Barros AJ, Wong KL, Johnson HL, Pariyo G, Franca GV, et al. Inequalities in full immunization coverage: trends in low- and middle-income countries. Bulletin of the World Health Organization. 2016;94(11):794–805b. https://doi.org/10.2471/blt.15.162172.

23. Nazroo JY. The structuring of ethnic inequalities in health: economic position, racial discrimination, and racism. American journal of public health. 2003;93(2):277–84. 10.2105/ajph.93.2.277.

24. Anderson I, Robson B, Connolly M, Al-Yaman F, Bjertness E, King A, et al. Indigenous and tribal peoples’ health (The Lancet-Lowitja Institute Global Collaboration): a population study. Lancet (London, England). 2016;388(10040):131–57. 10.1016/s0140-6736(16)00345-7.

25. Harper S, Lynch J. Methods for Measuring Cancer Disparities: Using Data Relevant to Healthy People 2010 Cancer-Related Objectives.. Bethesda, MD: Division of Cancer Control and Population Sciences of the National Cancer Institute; 2005.

26. Hosseinpoor AR, Bergen N, Barros AJ, Wong KL, Boerma T, Victora CG. Monitoring subnational regional inequalities in health: measurement approaches and challenges. International journal for equity in health. 2016;15(1):18. 10.1186/s12939-016-0307-y.

