## Supplementary materials for "Ethnic disparities in immunisation: analyses of zero-dose prevalence in 64 low- and middle-income countries"

**Supplementary material**

### Supplementary Table 1 – List of ethnic groups (or a proxy variable) and the grouping used in each country.

| **Country** | **Year** | **Ethnic group** | **Grouping (when applicable)** |
| --- | --- | --- | --- |
| Afghanistan | 2015 | Nuristani |  |
|  |  | Tajik |  |
|  |  | Uzbek |  |
|  |  | Hazara |  |
|  |  | Pashtun |  |
|  |  | Other |  |
|  |  | Turkmen |  |
|  |  | Pashai |  |
|  |  | Baloch |  |
| Angola | 2015 | Português |  |
|  |  | Chokwe/kioko |  |
|  |  | Umbundu |  |
|  |  | Nganguela |  |
|  |  | Nhaneca |  |
|  |  | Fiote | Grouped under "Other" category |
|  |  | Kikongo/ukongo |  |
|  |  | Kimbundu | Grouped under "Other" category |
|  |  | Kwanhama |  |
|  |  | Muhumbi |  |
|  |  | Luvale | Grouped under "Other" category |
|  |  | Other |  |
| Belize | 2015 | Mestizo/Spanish/Latino | Grouped under "Reference" category |
|  |  | Creole | Grouped under "African descent" category |
|  |  | Other | Grouped under "African descent" category |
|  |  | Maya | Grouped under "Indigenous" category |
|  |  | Garifuna | Grouped under "African descent" category |
| Benin | 2017 | Fon and related |  |
|  |  | Yoruba and related |  |
|  |  | Adja and related |  |
|  |  | Peulh and related |  |
|  |  | Other |  |
|  |  | Other |  |
|  |  | Dendi and related |  |
|  |  | Bariba and related |  |
|  |  | Betamaribe and related |  |
|  |  | Yoa, Lokpa and related |  |
| Bosnia_and_Herzegovina | 2011 | Bosnian |  |
|  |  | Serb |  |
|  |  | Croat |  |
| Burkina_Faso | 2010 | Mossi |  |
|  |  | Dagara |  |
|  |  | Bobo |  |
|  |  | Other |  |
|  |  | Sénoufo |  |
|  |  | Gourounsi |  |
|  |  | Fulfuldé / Peul |  |
|  |  | Gourmatché |  |
|  |  | Lobi |  |
|  |  | Bissa |  |
|  |  | Pays CEDEAO | Grouped under "Other" category |
|  |  | Dioula | Grouped under "Other" category |
|  |  | Touareg / Bella |  |
|  |  | Other | Grouped under "Other" category |
| Central African Republic | 2018 | Mandja |  |
|  |  | Gbaya |  |
|  |  | Sara |  |
|  |  | Banda |  |
|  |  | Haoussa |  |
|  |  | Ngbaka/Bantou |  |
|  |  | Yakoma/Sango |  |
|  |  | Other |  |
|  |  | Zandé/Nzakara | Grouped under "Other" category |
|  |  | Mboum |  |
| Cameroon | 2018 | Massa | Grouped under "Other" category |
|  |  | Guidar | Grouped under "Other" category |
|  |  | Mafa | Grouped under "Other" category |
|  |  | Gavar | Grouped under "Other" category |
|  |  | Bouwal | Grouped under "Other" category |
|  |  | Other |  |
|  |  | Boulou |  |
|  |  | Yambassa | Grouped under "Other" category |
|  |  | Ghomala-Cent | Grouped under "Other" category |
|  |  | Oku / Uku | Grouped under "Other" category |
|  |  | Bamoun |  |
|  |  | Bororo | Grouped under "Other" category |
|  |  | Haoussa | Grouped under "Other" category |
|  |  | Ghomala-Oues | Grouped under "Other" category |
|  |  | Foulbe |  |
|  |  | Nda'Nda-Sud | Grouped under "Other" category |
|  |  | Ewondo |  |
|  |  | Voute | Grouped under "Other" category |
|  |  | Africain-Voi | Grouped under "Other" category |
|  |  | Bassa | Grouped under "Other" category |
|  |  | Bansaw / Banso | Grouped under "Other" category |
|  |  | Tikar | Grouped under "Other" category |
|  |  | Bamileke |  |
|  |  | Ngomba | Grouped under "Other" category |
|  |  | Guiziga | Grouped under "Other" category |
|  |  | Kanuri | Grouped under "Other" category |
|  |  | Toupouri | Grouped under "Other" category |
|  |  | Daba | Grouped under "Other" category |
|  |  | Moundang | Grouped under "Other" category |
|  |  | Moussey | Grouped under "Other" category |
|  |  | Sara / Laka | Grouped under "Other" category |
|  |  | Maka / Makya | Grouped under "Other" category |
|  |  | Yanguere | Grouped under "Other" category |
|  |  | Other |  |
|  |  | Sanaga | Grouped under "Other" category |
|  |  | Manguissa | Grouped under "Other" category |
|  |  | Mambila | Grouped under "Other" category |
|  |  | Gbaya | Grouped under "Other" category |
|  |  | Banyang | Grouped under "Other" category |
|  |  | Bakossi | Grouped under "Other" category |
|  |  | Mambay | Grouped under "Other" category |
|  |  | Mada | Grouped under "Other" category |
|  |  | Kotoko | Grouped under "Other" category |
|  |  | Ngemba | Grouped under "Other" category |
|  |  | Mankon | Grouped under "Other" category |
|  |  | Bafia | Grouped under "Other" category |
|  |  | Arabe-Choa | Grouped under "Other" category |
|  |  | Bum | Grouped under "Other" category |
|  |  | Eton |  |
|  |  | Zime | Grouped under "Other" category |
|  |  | Mofou | Grouped under "Other" category |
|  |  | Fali | Grouped under "Other" category |
|  |  | Balom | Grouped under "Other" category |
|  |  | Africain-Aut | Grouped under "Other" category |
|  |  | Dschang-Cent | Grouped under "Other" category |
|  |  | Nda'Nda-Est | Grouped under "Other" category |
|  |  | Yamba | Grouped under "Other" category |
|  |  | Mbo | Grouped under "Other" category |
|  |  | Mousgoum | Grouped under "Other" category |
|  |  | Dourou | Grouped under "Other" category |
|  |  | Mboum | Grouped under "Other" category |
|  |  | Nkwen | Grouped under "Other" category |
|  |  | Ghomala-Sud | Grouped under "Other" category |
|  |  | Bamunka | Grouped under "Other" category |
|  |  | Eki | Grouped under "Other" category |
|  |  | Bafut | Grouped under "Other" category |
|  |  | Ngie | Grouped under "Other" category |
|  |  | Bambili | Grouped under "Other" category |
|  |  | Kozime | Grouped under "Other" category |
|  |  | Moghamo/Widi | Grouped under "Other" category |
|  |  | Ghomala-Nord | Grouped under "Other" category |
|  |  | Banen | Grouped under "Other" category |
|  |  | Kako | Grouped under "Other" category |
|  |  | Mouktele | Grouped under "Other" category |
|  |  | Fefe / Nufi-Ce | Grouped under "Other" category |
|  |  | Kom | Grouped under "Other" category |
|  |  | Ring | Grouped under "Other" category |
|  |  | Oshie/Ngishe | Grouped under "Other" category |
|  |  | Wimbum | Grouped under "Other" category |
|  |  | Noni | Grouped under "Other" category |
|  |  | Ntoumou | Grouped under "Other" category |
|  |  | Mvae | Grouped under "Other" category |
|  |  | Kwassio | Grouped under "Other" category |
|  |  | Kapsiki | Grouped under "Other" category |
|  |  | Zoulgo | Grouped under "Other" category |
|  |  | Podoko | Grouped under "Other" category |
|  |  | Metta | Grouped under "Other" category |
|  |  | Oroko | Grouped under "Other" category |
|  |  | Mungaka | Grouped under "Other" category |
|  |  | Medumba | Grouped under "Other" category |
|  |  | Mbam | Grouped under "Other" category |
|  |  | Lombe | Grouped under "Other" category |
|  |  | Mundum | Grouped under "Other" category |
|  |  | Bakwa | Grouped under "Other" category |
|  |  | Nda'Nda-Oues | Grouped under "Other" category |
|  |  | Yangben | Grouped under "Other" category |
|  |  | Fang | Grouped under "Other" category |
|  |  | Pinyin | Grouped under "Other" category |
|  |  | Bakweri | Grouped under "Other" category |
|  |  | Ngombale | Grouped under "Other" category |
|  |  | Pygmee | Grouped under "Other" category |
|  |  | Mbouko | Grouped under "Other" category |
|  |  | Bakoko | Grouped under "Other" category |
|  |  | Dschang-Oues | Grouped under "Other" category |
|  |  | Fong | Grouped under "Other" category |
|  |  | Fefe / Nufi-No | Grouped under "Other" category |
|  |  | Douala | Grouped under "Other" category |
|  |  | Ejagham | Grouped under "Other" category |
|  |  | Bafmeng | Grouped under "Other" category |
|  |  | Wandala | Grouped under "Other" category |
|  |  | Yebekolo | Grouped under "Other" category |
|  |  | Nchanti/Ncan | Grouped under "Other" category |
|  |  | Awing | Grouped under "Other" category |
|  |  | Kera | Grouped under "Other" category |
|  |  | Mbembe | Grouped under "Other" category |
|  |  | Pol/Pori | Grouped under "Other" category |
|  |  | Mezime / Mpo | Grouped under "Other" category |
|  |  | Nguiembong | Grouped under "Other" category |
|  |  | Mouyang | Grouped under "Other" category |
|  |  | Aghem | Grouped under "Other" category |
|  |  | Koutine | Grouped under "Other" category |
|  |  | Mvele | Grouped under "Other" category |
|  |  | Momo | Grouped under "Other" category |
|  |  | Kaka | Grouped under "Other" category |
|  |  | Bandem | Grouped under "Other" category |
|  |  | Kobotchi | Grouped under "Other" category |
|  |  | Fefe / Nufi | Grouped under "Other" category |
|  |  | Megaka / Bagam | Grouped under "Other" category |
|  |  | Djanti | Grouped under "Other" category |
| Chad | 2014 | Massa/Mousseye/Mousgoume |  |
|  |  | Ouadaï/Maba/Massalit/Mimi |  |
|  |  | Other |  |
|  |  | Sara |  |
|  |  | Peul/Foulbé/Bodoré |  |
|  |  | Dadajo/Kibet/Mouro |  |
|  |  | Gabri/Kabalaye/Nangtchéré/Soumraye | Grouped under "Other" category |
|  |  | Arab |  |
|  |  | Kanembou/Bornou/Boudouma |  |
|  |  | Other |  |
|  |  | Boulala/Médégo/Kouka |  |
|  |  | Zaghawa/Bideyat/Kobé |  |
|  |  | Gorane |  |
|  |  | Tama / Assongori / Mararit |  |
|  |  | Other |  |
|  |  | Toupouri/Kéra |  |
|  |  | Baguirmi/Barma | Grouped under "Other" category |
|  |  | Marba/Lélé/Mesmé |  |
|  |  | Mesmedjé/Massalat/Kadjaksé | Grouped under "Other" category |
|  |  | Moundang |  |
|  |  | Bidio/Migami/Kenga/Dangléat |  |
|  |  | Karo/Zimé/Pévé |  |
| Colombia | 2010 | Native Colombian | Grouped under "Indigenous" category |
|  |  | Other | Grouped under "Reference" category |
|  |  | Black/Mulato/Afro-Colombian/Afro-Descendent | Grouped under "African descent" category |
|  |  | Palanquero From San Basilio | Grouped under "African descent" category |
|  |  | Raizal From Archipelago (San Andres) | Grouped under "African descent" category |
| Congo_Brazzaville | 2014 | Other |  |
|  |  | Kongo |  |
|  |  | Eshira | Grouped under "Other" category |
|  |  | Other |  |
|  |  | Mbosi |  |
|  |  | Tekes |  |
|  |  | Mbetis |  |
|  |  | Sangha-Likouala |  |
| Congo_Democratic_Republic | 2017 | Bantou |  |
|  |  | Other |  |
|  |  | Pygmees | Grouped under "Other" category |
|  |  | Nilotique |  |
|  |  | Soudanais |  |
| Costa_Rica | 2018 | Ninguna | Grouped under "Reference" category |
|  |  | Blanco | Grouped under "Reference" category |
|  |  | Negro/afrodescendiente | Grouped under "African descent" category |
|  |  | Mestizo | Grouped under "African descent" category |
|  |  | Mulato | Grouped under "African descent" category |
|  |  | Otra | Grouped under "Reference" category |
|  |  | Indígena | Grouped under "Indigenous" category |
|  |  | Chino | Grouped under "Reference" category |
| Cote_dIvoire | 2016 | Gur |  |
|  |  | Mandé du Sud |  |
|  |  | Mandé du Nord |  |
|  |  | Other |  |
|  |  | Other |  |
|  |  | Akan |  |
|  |  | Krou |  |
| Cuba | 2019 | Mulato, mestizo / Otro | Grouped under "African descent" category |
|  |  | Blanco | Grouped under "Reference" category |
|  |  | Negro | Grouped under "African descent" category |
| Dominican_Republic | 2014 | Español | Grouped under "Reference" category |
|  |  | Creole | Grouped under "African descent" category |
|  |  | Otra | Grouped under "Reference" category |
| Ethiopia | 2016 | Nuwer |  |
|  |  | Welaita | Grouped under "Other" category |
|  |  | Affar |  |
|  |  | Oromo |  |
|  |  | Amhara |  |
|  |  | Tigrie |  |
|  |  | Hadiya | Grouped under "Other" category |
|  |  | Somalie |  |
|  |  | Guragie |  |
|  |  | Kembata | Grouped under "Other" category |
|  |  | Silte | Grouped under "Other" category |
|  |  | Me'Enite | Grouped under "Other" category |
|  |  | Bench | Grouped under "Other" category |
|  |  | Berta |  |
|  |  | Gedeo | Grouped under "Other" category |
|  |  | Sidama | Grouped under "Other" category |
|  |  | Goffa | Grouped under "Other" category |
|  |  | Agew-Awi | Grouped under "Other" category |
|  |  | Anyiwak | Grouped under "Other" category |
|  |  | Other |  |
|  |  | Gamo | Grouped under "Other" category |
|  |  | Sheko | Grouped under "Other" category |
|  |  | Mejenger | Grouped under "Other" category |
|  |  | Bena | Grouped under "Other" category |
|  |  | Kefficho | Grouped under "Other" category |
|  |  | Yem | Grouped under "Other" category |
|  |  | Gumuz | Grouped under "Other" category |
|  |  | Shinasha | Grouped under "Other" category |
|  |  | Agew Hamyra | Grouped under "Other" category |
|  |  | Mao | Grouped under "Other" category |
|  |  | Komo | Grouped under "Other" category |
|  |  | Burji | Grouped under "Other" category |
|  |  | Ari | Grouped under "Other" category |
|  |  | Dawuro | Grouped under "Other" category |
|  |  | Harari | Grouped under "Other" category |
|  |  | Other | Grouped under "Other" category |
|  |  | Argoba | Grouped under "Other" category |
|  |  | Dizi | Grouped under "Other" category |
|  |  | Konta | Grouped under "Other" category |
|  |  | Derashe | Grouped under "Other" category |
| Gabon | 2012 | Shira-Punu/Vili |  |
|  |  | Fang |  |
|  |  | Kota-Kele |  |
|  |  | Other | Grouped under "Other" category |
|  |  | Okande-Tsogho | Grouped under "Other" category |
|  |  | Nzabi-Duma |  |
|  |  | Pygmee | Grouped under "Other" category |
|  |  | Mbede-Teke |  |
|  |  | Myene | Grouped under "Other" category |
| Gambia | 2018 | Fula |  |
|  |  | Mandinka |  |
|  |  | Sarahule |  |
|  |  | Other | Grouped under "Other" category |
|  |  | Wollof |  |
|  |  | Other | Grouped under "Other" category |
|  |  | Jola |  |
| Ghana | 2017 | Gruma |  |
|  |  | Mole Dagbani |  |
|  |  | Grusi |  |
|  |  | Akan |  |
|  |  | Other | Grouped under "Other" category |
|  |  | Ga/Damgme |  |
|  |  | Guan |  |
|  |  | Ewe |  |
|  |  | Mande | Grouped under "Other" category |
| Guatemala | 2014 | Ladina/Mestiza | Grouped under "Reference" category |
|  |  | Maya | Grouped under "Indigenous" category |
|  |  | Xinca | Grouped under "Indigenous" category |
|  |  | Garífuna | Grouped under "African descent" category |
| Guinea | 2018 | Peulh |  |
|  |  | Soussou |  |
|  |  | Malinké |  |
|  |  | Guerzé |  |
|  |  | Kissi |  |
| Guinea_Bissau | 2018 | Balanta |  |
|  |  | Beafada |  |
|  |  | Other | Grouped under "Other" category |
|  |  | Mandinga |  |
|  |  | Papel |  |
|  |  | Fula |  |
|  |  | Mancanha | Grouped under "Other" category |
|  |  | Manjaco |  |
|  |  | Felupe | Grouped under "Other" category |
| Guyana | 2014 | Mixed Race | Grouped under "Reference" category |
|  |  | East Indian | Grouped under "Reference" category |
|  |  | African | Grouped under "African descent" category |
|  |  | Amerindian | Grouped under "Indigenous" category |
| Honduras | 2011 | Garifuna | Grouped under "African descent" category |
|  |  | Lenca | Grouped under "Indigenous" category |
|  |  | DK/none | Grouped under "Reference" category |
|  |  | Misquito | Grouped under "Indigenous" category |
|  |  | Tolupán | Grouped under "Indigenous" category |
|  |  | Other | Grouped under "Reference" category |
|  |  | Negro inglés | Grouped under "African descent" category |
|  |  | Maya chorti | Grouped under "Indigenous" category |
|  |  | Tawaka (sumo) | Grouped under "Indigenous" category |
|  |  | Pech (paya) | Grouped under "Indigenous" category |
| India | 2015 | Caste |  |
|  |  | Tribe |  |
|  |  | No caste/Tribe |  |
| Iraq | 2018 | Arabic |  |
|  |  | Kurdish |  |
| Jordan | 2017 | Jordanian |  |
|  |  | Egyptian | Grouped under "Other" category |
|  |  | Syrian |  |
|  |  | Other | Grouped under "Other" category |
|  |  | Iraqi |  |
|  |  | Other | Grouped under "Other" category |
| Kazakhstan | 2015 | Kazakh |  |
|  |  | Other | Grouped under "Other" category |
|  |  | Russian |  |
| Kenya | 2014 | Luo |  |
|  |  | Luhya |  |
|  |  | Iteso | Grouped under "Other" category |
|  |  | Taita/Taveta | Grouped under "Other" category |
|  |  | Other | Grouped under "Other" category |
|  |  | Kuria | Grouped under "Other" category |
|  |  | Kikuya |  |
|  |  | Kalenjin |  |
|  |  | Kamba |  |
|  |  | Somali |  |
|  |  | Turkana |  |
|  |  | Kisii |  |
|  |  | Mijikenda/Swahili |  |
|  |  | Meru |  |
|  |  | Maasai |  |
|  |  | Boran |  |
|  |  | Samburu |  |
|  |  | Mbere | Grouped under "Other" category |
|  |  | Embu | Grouped under "Other" category |
|  |  | Rendille | Grouped under "Other" category |
|  |  | Orma | Grouped under "Other" category |
|  |  | Pokomo |  |
|  |  | Gabbra | Grouped under "Other" category |
| Kyrgyzstan | 2018 | Uzbek |  |
|  |  | Kyrgyz |  |
| Laos | 2017 | Chinese-Tibetan |  |
|  |  | Hmong-Mien |  |
|  |  | Lao-Tai |  |
|  |  | Mon-Khmer |  |
| Malawi | 2015 | Yao |  |
|  |  | Ngoni |  |
|  |  | Chewa |  |
|  |  | Lomwe |  |
|  |  | Sena |  |
|  |  | Other | Grouped under "Other" category |
|  |  | Tumbuka |  |
|  |  | Nyanga | Grouped under "Other" category |
|  |  | Nkhonde | Grouped under "Other" category |
|  |  | Tonga |  |
|  |  | Mang'Anja |  |
| Mali | 2018 | Peulh |  |
|  |  | Sonraï |  |
|  |  | Bambara |  |
|  |  | Touareg/Bella |  |
|  |  | Other | Grouped under "Other" category |
|  |  | Sénoufo/Minianka |  |
|  |  | Dogon |  |
|  |  | Sarakole/Soninke/Marka |  |
|  |  | Malinke |  |
|  |  | Bobo | Grouped under "Other" category |
|  |  | Other | Grouped under "Other" category |
|  |  | Ecowas Countries | Grouped under "Other" category |
|  |  | Other | Grouped under "Other" category |
| Mauritania | 2015 | Arabe |  |
|  |  | Soninké |  |
|  |  | Poular |  |
|  |  | Wolof | Grouped under "Other" category |
|  |  | Other | Grouped under "Other" category |
| Mexico | 2015 | Hogar no indígena | Grouped under "Reference" category |
|  |  | Hogar indígena | Grouped under "Indigenous" category |
| Mongolia | 2018 | Khalkh |  |
|  |  | Other | Grouped under "Other" category |
|  |  | Kazakh |  |
| Montenegro | 2013 | Montenegrin |  |
|  |  | Serbian |  |
|  |  | Albanian | Grouped under "Other" category |
|  |  | Roma | Grouped under "Other" category |
|  |  | Muslim | Grouped under "Other" category |
|  |  | Other | Grouped under "Other" category |
|  |  | Croat | Grouped under "Other" category |
|  |  | Bosniak | Grouped under "Other" category |
| Mozambique | 2015 | Cisena |  |
|  |  | Elomwe | Grouped under "Other" category |
|  |  | Other | Grouped under "Other" category |
|  |  | Xitswa |  |
|  |  | Português |  |
|  |  | Xichangana |  |
|  |  | Ciyao | Grouped under "Other" category |
|  |  | Emakhuwa |  |
|  |  | Cinyanja |  |
|  |  | Cinyungwe | Grouped under "Other" category |
|  |  | Shona | Grouped under "Other" category |
|  |  | Echuwabo | Grouped under "Other" category |
|  |  | Cindau |  |
| Myanmar | 2015 | Myanmar |  |
|  |  | Other | Grouped under "Other" category |
| Namibia | 2013 | San | Grouped under "Other" category |
|  |  | Oshiwambo |  |
|  |  | Damara/Nama |  |
|  |  | Afrikaans |  |
|  |  | Lozi |  |
|  |  | Kwangali |  |
|  |  | English | Grouped under "Other" category |
|  |  | Other | Grouped under "Other" category |
|  |  | Herero |  |
| Nepal | 2019 | Kami |  |
|  |  | Chhetree |  |
|  |  | Tamang |  |
|  |  | Lohar | Grouped under "Other" category |
|  |  | Tharu |  |
|  |  | Musalman |  |
|  |  | Kewat | Grouped under "Other" category |
|  |  | Brahman - Hill |  |
|  |  | Magar |  |
|  |  | Thakuri | Grouped under "Other" category |
|  |  | Dhobi | Grouped under "Other" category |
|  |  | Yadav | Grouped under "Other" category |
|  |  | Other | Grouped under "Other" category |
|  |  | Kurmi | Grouped under "Other" category |
|  |  | Koiri/Kushwaha | Grouped under "Other" category |
|  |  | Dusadh/Pasawan/Pasi | Grouped under "Other" category |
|  |  | Kalwar | Grouped under "Other" category |
|  |  | Chamar/Harijan/Ram | Grouped under "Other" category |
|  |  | Kumal | Grouped under "Other" category |
|  |  | Kathbaniyan | Grouped under "Other" category |
|  |  | Damai/Dholi | Grouped under "Other" category |
|  |  | Newar | Grouped under "Other" category |
|  |  | Sarki | Grouped under "Other" category |
|  |  | Other | Grouped under "Other" category |
|  |  | Rajbhar | Grouped under "Other" category |
|  |  | Hajam/Thakur | Grouped under "Other" category |
|  |  | Musahar | Grouped under "Other" category |
|  |  | Gurung | Grouped under "Other" category |
|  |  | Chepang/Praja | Grouped under "Other" category |
|  |  | Darai | Grouped under "Other" category |
|  |  | Gharti/Bhujel | Grouped under "Other" category |
|  |  | Other | Grouped under "Other" category |
|  |  | Rai | Grouped under "Other" category |
|  |  | Other | Grouped under "Other" category |
|  |  | Thakali | Grouped under "Other" category |
|  |  | Majhi | Grouped under "Other" category |
|  |  | Sunuwar | Grouped under "Other" category |
|  |  | Limbu | Grouped under "Other" category |
|  |  | Sanyasi/Dashnami | Grouped under "Other" category |
|  |  | Tajpuriya | Grouped under "Other" category |
|  |  | Ghale | Grouped under "Other" category |
|  |  | Pahari | Grouped under "Other" category |
|  |  | Danuwar | Grouped under "Other" category |
|  |  | Sonar | Grouped under "Other" category |
|  |  | Mallaha | Grouped under "Other" category |
|  |  | Bin | Grouped under "Other" category |
|  |  | Tatma/Tatwa | Grouped under "Other" category |
|  |  | Rajput | Grouped under "Other" category |
|  |  | Kumhar | Grouped under "Other" category |
|  |  | Teli | Grouped under "Other" category |
|  |  | Brahman - Tarai | Grouped under "Other" category |
|  |  | Dhanuk | Grouped under "Other" category |
|  |  | Kori | Grouped under "Other" category |
|  |  | Dev | Grouped under "Other" category |
|  |  | Baraee | Grouped under "Other" category |
|  |  | Kanu | Grouped under "Other" category |
|  |  | Nuniya | Grouped under "Other" category |
|  |  | Mali | Grouped under "Other" category |
|  |  | Gaderi/Bhedhar | Grouped under "Other" category |
|  |  | Bantar/Sardar | Grouped under "Other" category |
|  |  | Amat | Grouped under "Other" category |
|  |  | Bahing | Grouped under "Other" category |
|  |  | Marwadi | Grouped under "Other" category |
|  |  | Bangali | Grouped under "Other" category |
|  |  | Gangai | Grouped under "Other" category |
|  |  | Haluwai | Grouped under "Other" category |
|  |  | Yakkha | Grouped under "Other" category |
|  |  | Rajbansi | Grouped under "Other" category |
|  |  | Aathpariya | Grouped under "Other" category |
|  |  | Jhangad/Dhagar | Grouped under "Other" category |
|  |  | Satar/Santhal | Grouped under "Other" category |
|  |  | Sherpa | Grouped under "Other" category |
| Niger | 2012 | Haoussa |  |
|  |  | Zarma |  |
|  |  | Kanouri/Toubou |  |
|  |  | French | Grouped under "Other" category |
|  |  | Tamasheq | Grouped under "Other" category |
|  |  | Fulfulde | Grouped under "Other" category |
|  |  | Other | Grouped under "Other" category |
|  |  | Gourmantchéma | Grouped under "Other" category |
|  |  | Arab | Grouped under "Other" category |
| Nigeria | 2018 | Hausa |  |
|  |  | Fulani |  |
|  |  | Kanuri/Beriberi |  |
|  |  | Other | Grouped under "Other" category |
|  |  | Yoruba |  |
|  |  | Ekoi | Grouped under "Other" category |
|  |  | Igbo |  |
|  |  | Ibibio |  |
|  |  | Tiv |  |
|  |  | Ijaw/Izon |  |
|  |  | Igala |  |
| North_Macedonia | 2018 | Macedonian |  |
|  |  | Albanian |  |
| Pakistan | 2017 | Urdu |  |
|  |  | Punjabi |  |
|  |  | Pushto |  |
|  |  | Sariaki |  |
|  |  | Other | Grouped under "Other" category |
|  |  | English | Grouped under "Other" category |
|  |  | Baluchi |  |
|  |  | Sindhi |  |
| Panama | 2013 | Otro grupo | Grouped under "Reference" category |
|  |  | Negro o afrodescendiente | Grouped under "African descent" category |
|  |  | Indígena | Grouped under "Indigenous" category |
| Papua_New_Guinea | 2016 | Tok Ples |  |
|  |  | Pidgin |  |
|  |  | English |  |
| Paraguay | 2016 | Indígena | Grouped under "Indigenous" category |
|  |  | Hablante guaraní y castellano | Grouped under "Reference" category |
|  |  | Hablante sólo guaraní | Grouped under "Indigenous" category |
|  |  | Hablante sólo castellano | Grouped under "Reference" category |
|  |  | Hablante de otro idioma | Grouped under "Reference" category |
| Peru | 2019 | Otra lengua nativa u originaria | Grouped under "Indigenous" category |
|  |  | Shipibo/Konibo | Grouped under "Indigenous" category |
|  |  | Castellano | Grouped under "Reference" category |
|  |  | Quechua | Grouped under "Indigenous" category |
|  |  | Achuar | Grouped under "Indigenous" category |
|  |  | Portugués | Grouped under "Reference" category |
|  |  | Awajún/Aguaruna | Grouped under "Indigenous" category |
|  |  | Shawi/Chayahuita | Grouped under "Indigenous" category |
|  |  | Matsigenka/ Machiguenga | Grouped under "Indigenous" category |
|  |  | Aimara | Grouped under "Indigenous" category |
|  |  | Ashaninka | Grouped under "Indigenous" category |
| Philippines | 2017 | Ilonggo |  |
|  |  | Other | Grouped under "Other" category |
|  |  | Tausog | Grouped under "Other" category |
|  |  | Cebuano |  |
|  |  | Visaya |  |
|  |  | Maranao |  |
|  |  | Ilokano |  |
|  |  | Waray |  |
|  |  | Tagalog |  |
|  |  | Bikolano |  |
|  |  | Kapampangan | Grouped under "Other" category |
| Senegal | 2019 | Wolof |  |
|  |  | Poular |  |
|  |  | Other | Grouped under "Other" category |
|  |  | Mandingue/ Socé |  |
|  |  | Serer |  |
|  |  | Not Senegalese | Grouped under "Other" category |
|  |  | Diola | Grouped under "Other" category |
|  |  | Soninké | Grouped under "Other" category |
| Sierra_Leone | 2019 | Mende |  |
|  |  | Temne |  |
|  |  | Loko | Grouped under "Other" category |
|  |  | Limba |  |
|  |  | Fullah |  |
|  |  | Other |  |
|  |  | Korankoh |  |
|  |  | Sherbro | Grouped under "Other" category |
|  |  | Kono |  |
|  |  | Mandingo | Grouped under "Other" category |
|  |  | Other | Grouped under "Other" category |
|  |  | Creole | Grouped under "Other" category |
| South_Africa | 2016 | Black/African |  |
|  |  | Coloured |  |
| Suriname | 2018 | Hindustani | Grouped under "Reference" category |
|  |  | Indigenous/Amerindian | Grouped under "Indigenous" category |
|  |  | Maroon | Grouped under "African descent" category |
|  |  | Creole | Grouped under "Reference" category |
|  |  | Mixed ethnicity | Grouped under "Reference" category |
|  |  | Javanese | Grouped under "Reference" category |
|  |  | Other | Grouped under "Reference" category |
| Tajikistan | 2017 | Other | Grouped under "Other" category |
|  |  | Russian |  |
|  |  | Tajik | Grouped under "Other" category |
| Thailand | 2019 | Thai |  |
|  |  | Non-Thai |  |
| Timor_Leste | 2016 | Tetum |  |
|  |  | Other | Grouped under "Other" category |
|  |  | Bahasa | Grouped under "Other" category |
| Togo | 2017 | Akposso/Akébou |  |
|  |  | Other | Grouped under "Other" category |
|  |  | Kabye-Tem |  |
|  |  | Other | Grouped under "Other" category |
|  |  | Adja-Ewe |  |
|  |  | Ana-Ife | Grouped under "Other" category |
|  |  | Paragourma | Grouped under "Other" category |
| Turkmenistan | 2015 | Turkmen |  |
|  |  | Uzbek |  |
| Uganda | 2016 | Acholi |  |
|  |  | Iteso |  |
|  |  | Lango |  |
|  |  | Madi | Grouped under "Other" category |
|  |  | Kuku | Grouped under "Other" category |
|  |  | Kakwa | Grouped under "Other" category |
|  |  | Aringa | Grouped under "Other" category |
|  |  | Bakonzo |  |
|  |  | Lugbara |  |
|  |  | Banyoro |  |
|  |  | Alur |  |
|  |  | Jonam | Grouped under "Other" category |
|  |  | Kebu (Okebu) | Grouped under "Other" category |
|  |  | Nubi | Grouped under "Other" category |
|  |  | Other | Grouped under "Other" category |
|  |  | Bagisu |  |
|  |  | Chope | Grouped under "Other" category |
|  |  | Baruli | Grouped under "Other" category |
|  |  | Basoga |  |
|  |  | Banyankore |  |
|  |  | Bagwere |  |
|  |  | Baganda |  |
|  |  | Bagungu | Grouped under "Other" category |
|  |  | Bakiga |  |
|  |  | Bafumbira |  |
|  |  | Banyarwanda |  |
|  |  | Batoro |  |
|  |  | Bahororo | Grouped under "Other" category |
|  |  | Batagwenda | Grouped under "Other" category |
|  |  | Babwisi | Grouped under "Other" category |
|  |  | Baamba | Grouped under "Other" category |
|  |  | Batuku | Grouped under "Other" category |
|  |  | Banyaruguru | Grouped under "Other" category |
|  |  | Banyole |  |
|  |  | Bakenyi | Grouped under "Other" category |
|  |  | Jopadhola |  |
|  |  | Karimojong |  |
|  |  | Basamia | Grouped under "Other" category |
|  |  | Barundi | Grouped under "Other" category |
|  |  | Pokot | Grouped under "Other" category |
|  |  | Jie | Grouped under "Other" category |
|  |  | Ik (Teuso) | Grouped under "Other" category |
|  |  | Sabiny | Grouped under "Other" category |
|  |  | Ethur | Grouped under "Other" category |
|  |  | Dodoth | Grouped under "Other" category |
|  |  | Mening | Grouped under "Other" category |
|  |  | Napore | Grouped under "Other" category |
|  |  | Kumam | Grouped under "Other" category |
|  |  | Banyara | Grouped under "Other" category |
| Vietnam | 2013 | Kinh |  |
|  |  | Non-Kinh |  |
| Zambia | 2018 | Lozi |  |
|  |  | Tonga |  |
|  |  | Bemba |  |
|  |  | Kaonde |  |
|  |  | Nyanja |  |
|  |  | Other | Grouped under "Other" category |
|  |  | English | Grouped under "Other" category |
|  |  | Lunda |  |
|  |  | Luvale |  |
| Zimbabwe | 2019 | Other | Grouped under "Other" category |
|  |  | Shona |  |
|  |  | Ndebele |  |
|  |  | English | Grouped under "Other" category |

### Supplementary Table 2 - Countries included in the analysis, number of ethnic groups by country and no-DPT prevalence and prevalence ratio for ethnic groups.

| **Country** | **Year** | **Source** | **Gavi eligibility** | **Variable** | **Number of children** | **Ethnic groups** | **No-DPT prevalence by ethnic group** | | | | | **Ratio** |
| --- | --- | --- | --- | --- | --- | --- | --- | --- | --- | --- | --- | --- |
|  |  |  |  |  |  |  | **Median** | **Lowest** | **95%CI** | **Highest** | **95%CI** |  |
| Afghanistan | 2015 | DHS | Yes | Ethnicity | 5809 | 9 | 32 | 15 | 6;23 | 99 | 97;100 | 6·7 |
| Angola | 2015 | DHS | Yes | Language | 2845 | 9 | 43 | 16 | 9;23 | 66 | 47;86 | 4·2 |
| Belize | 2015 | MICS | No | Ethnicity | 503 | 3 | 11 | 4 | 1;7 | 13 | 5;21 | 3·4 |
| Benin | 2017 | DHS | Yes | Ethnicity | 2522 | 9 | 14 | 8 | 6;10 | 51 | 42;60 | 6·3 |
| Bosnia and Herzegovina | 2011 | MICS | No | Language | 512 | 3 | 3 | 2 | 0;4 | 3 | 2;5 | 1·7 |
| Burkina Faso | 2010 | DHS | Yes | Ethnicity | 2784 | 11 | 4 | 0 | 0;0 | 32 | 14;49 | - |
| Cameroon | 2018 | DHS | Yes | Ethnicity | 1824 | 7 | 10 | 8 | 3;14 | 28 | 19;37 | 3·3 |
| Central African Republic | 2018 | MICS | Yes | Ethnicity | 1688 | 9 | 44 | 34 | 23;46 | 58 | 46;71 | 1·7 |
| Chad | 2014 | DHS | Yes | Ethnicity | 2810 | 17 | 43 | 8 | 3;13 | 79 | 66;92 | 9·5 |
| Colombia | 2010 | DHS | No | Ethnicity | 3434 | 3 | 3 | 3 | 1;5 | 4 | 2;7 | 1·4 |
| Congo Brazzaville | 2014 | MICS | Yes | Ethnicity | 1772 | 6 | 22 | 8 | 5;10 | 29 | 19;39 | 3·7 |
| Congo, DR | 2017 | MICS | Yes | Ethnicity | 4222 | 3 | 34 | 20 | 8;32 | 36 | 27;46 | 1·8 |
| Costa Rica | 2018 | MICS | No | Ethnicity | 625 | 2 | 3 | 1 | 0;2 | 5 | 1;8 | 5·0 |
| Côte d'Ivoire | 2016 | MICS | Yes | Ethnicity | 1783 | 6 | 22 | 10 | 7;13 | 25 | 20;31 | 2·6 |
| Cuba | 2019 | MICS | No | Skin color | 1119 | 2 | 3 | 1 | 1;2 | 5 | 0;10 | 3·6 |
| Dominican Republic | 2014 | MICS | No | Language | 3917 | 2 | 15 | 8 | 6;9 | 22 | 16;27 | 2·9 |
| Ethiopia | 2016 | DHS | Yes | Ethnicity | 1926 | 9 | 26 | 6 | 2;11 | 55 | 29;82 | 9·1 |
| Gabon | 2012 | DHS | No | Ethnicity | 1073 | 6 | 12 | 9 | 3;15 | 15 | 9;22 | 1·7 |
| Gambia | 2018 | MICS | Yes | Ethnicity | 1895 | 6 | 3 | 1 | 0;2 | 6 | 2;9 | 4·8 |
| Ghana | 2017 | MICS | Yes | Ethnicity | 1680 | 8 | 4 | 2 | 0;5 | 7 | 1;13 | 3·3 |
| Guatemala | 2014 | DHS | No | Ethnicity | 2401 | 2 | 3 | 1 | 1;2 | 4 | 3;5 | 2·9 |
| Guinea | 2018 | DHS | Yes | Ethnicity | 1390 | 5 | 31 | 15 | 6;25 | 54 | 48;59 | 3·5 |
| Guinea Bissau | 2018 | MICS | Yes | Ethnicity | 1409 | 7 | 6 | 1 | 0;3 | 11 | 1;21 | 9·6 |
| Guyana | 2014 | MICS | Yes | Ethnicity | 686 | 3 | 4 | 4 | 1;7 | 6 | 2;10 | 1·6 |
| Honduras | 2011 | DHS | Yes | Ethnicity | 2275 | 3 | 1 | 0 | 0;0 | 1 | 1;2 | 10·0 |
| India | 2015 | DHS | Yes | Caste | 48932 | 3 | 11 | 10 | 10;11 | 14 | 13;16 | 1·4 |
| Iraq | 2018 | MICS | No | Language | 3160 | 2 | 11 | 7 | 3;11 | 15 | 12;17 | 2·2 |
| Jordan | 2017 | DHS | No | Ethnicity | 1945 | 3 | 11 | 6 | 5;8 | 20 | 3;36 | 3·0 |
| Kazakhstan | 2015 | MICS | No | Ethnicity | 1103 | 3 | 4 | 3 | 2;4 | 13 | 5;21 | 4·3 |
| Kenya | 2014 | DHS | Yes | Ethnicity | 4051 | 15 | 2 | 0 | 0;1 | 11 | 7;16 | 27·5 |
| Kyrgyzstan | 2018 | MICS | Yes | Ethnicity | 602 | 2 | 11 | 8 | 5;11 | 14 | 5;22 | 1·7 |
| Laos | 2017 | MICS | Yes | Ethnicity | 2194 | 4 | 35 | 19 | 16;22 | 43 | 37;49 | 2·3 |
| Malawi | 2015 | DHS | Yes | Ethnicity | 3248 | 9 | 2 | 1 | 0;3 | 7 | 2;12 | 7·9 |
| Mali | 2018 | DHS | Yes | Ethnicity | 1946 | 9 | 21 | 8 | 4;13 | 52 | 32;71 | 6·3 |
| Mauritania | 2015 | MICS | Yes | Language | 2088 | 3 | 15 | 15 | 12;17 | 25 | 9;41 | 1·7 |
| Mexico | 2015 | MICS | No | Ethnicity | 1536 | 2 | 9 | 8 | 5;10 | 11 | 4;17 | 1·4 |
| Mongolia | 2018 | MICS | No | Ethnicity | 1070 | 3 | 3 | 2 | 0;4 | 12 | 4;20 | 6·7 |
| Montenegro | 2013 | MICS | No | Ethnicity | 249 | 3 | 7 | 4 | 0;7 | 11 | 0;22 | 3·0 |
| Mozambique | 2015 | DHS | Yes | Language | 1026 | 8 | 11 | 1 | 0;3 | 27 | 9;45 | 24·4 |
| Myanmar | 2015 | DHS | Yes | Language | 915 | 2 | 18 | 11 | 8;14 | 26 | 15;37 | 2·4 |
| Namibia | 2013 | DHS | No | Language | 990 | 7 | 7 | 6 | 3;8 | 19 | 3;35 | 3·4 |
| Nepal | 2019 | MICS | Yes | Ethnicity | 1327 | 8 | 8 | 4 | 0;9 | 21 | 9;34 | 4·9 |
| Niger | 2012 | DHS | Yes | Language | 2151 | 4 | 20 | 5 | 3;8 | 40 | 29;50 | 7·9 |
| Nigeria | 2018 | DHS | Yes | Ethnicity | 6057 | 10 | 26 | 6 | 4;8 | 64 | 58;69 | 10·1 |
| North Macedonia | 2018 | MICS | No | Ethnicity | 272 | 2 | 2 | 2 | 0;3 | 3 | 0;9 | 1·8 |
| Pakistan | 2017 | DHS | Yes | Language | 2314 | 7 | 17 | 1 | 0;2 | 28 | 21;35 | 31·4 |
| Panama | 2013 | MICS | No | Ethnicity | 1323 | 3 | 6 | 5 | 1;9 | 14 | 8;19 | 2·8 |
| Papua New Guinea | 2016 | DHS | Yes | Language | 1709 | 3 | 36 | 15 | 1;29 | 38 | 32;45 | 2·6 |
| Paraguay | 2016 | MICS | No | Ethnicity | 1012 | 2 | 5 | 4 | 2;6 | 7 | 4;9 | 1·7 |
| Peru | 2019 | DHS | No | Ethnicity | 4201 | 2 | 6 | 5 | 4;6 | 7 | 4;10 | 1·5 |
| Philippines | 2017 | DHS | No | Ethnicity | 1986 | 9 | 13 | 4 | 1;7 | 64 | 49;79 | 16·4 |
| Senegal | 2019 | DHS | Yes | Ethnicity | 1183 | 5 | 3 | 2 | 0;4 | 6 | 2;9 | 2·7 |
| Sierra Leone | 2019 | DHS | Yes | Ethnicity | 1861 | 7 | 6 | 0 | 0;0 | 9 | 0;18 | - |
| South Africa | 2016 | DHS | No | Ethnicity | 660 | 2 | 8 | 8 | 1;15 | 9 | 6;12 | 1·2 |
| Suriname | 2018 | MICS | No | Ethnicity | 719 | 2 | 20 | 18 | 12;24 | 23 | 15;30 | 1·2 |
| Tajikistan | 2017 | DHS | Yes | Language | 1297 | 2 | 7 | 6 | 2;11 | 8 | 6;10 | 1·3 |
| Thailand | 2019 | MICS | No | Language | 2879 | 2 | 9 | 2 | 1;3 | 15 | 2;29 | 7·6 |
| Timor-Leste | 2016 | DHS | Yes | Language | 1423 | 2 | 19 | 15 | 4;26 | 22 | 19;25 | 1·4 |
| Togo | 2017 | MICS | Yes | Ethnicity | 973 | 4 | 9 | 6 | 2;10 | 12 | 4;21 | 2·3 |
| Turkmenistan | 2015 | MICS | No | Language | 752 | 2 | 0 | 0 | 0;0 | 1 | 0;1 | - |
| Uganda | 2016 | DHS | Yes | Ethnicity | 2922 | 20 | 4 | 1 | 0;2 | 15 | 1;30 | 30·4 |
| Vietnam | 2013 | MICS | Yes | Ethnicity | 785 | 2 | 7 | 2 | 1;4 | 12 | 6;18 | 5·0 |
| Zambia | 2018 | DHS | Yes | Language | 1928 | 8 | 2 | 1 | 0;2 | 10 | 0;23 | 17·2 |
| Zimbabwe | 2019 | MICS | Yes | Language | 1153 | 3 | 2 | 0 | 0;0 | 6 | 4;8 | - |

Legend: no-DPT: children who did not received any doses of the diphtheria-tetanus-pertussis-containing vaccine. CI: confidence interval. Lowest refers to the ethnic group with the lowest no-DPT prevalence in the country. Highest refers to the ethnic group with the highest no-DPT prevalence in the country.

Ratio, as a relative measure, is influenced by low values, therefore the highest ratios are probably due to the low no-DPT prevalence in the lowest group and might be influenced by rounding in no-DPT prevalence in the lowest and highest groups.

### Supplementary Table 3 – Crude and Adjusted no-DPT prevalence by ethnic group.

|  |  |  | **Crude** | | | | **Adjusted** | | | |
| --- | --- | --- | --- | --- | --- | --- | --- | --- | --- | --- |
| **Country** | **Year** | **Ethnic group** | **No-DPT prevalence** | **95%CI** | | **p value** | **No-DPT prevalence** | **95%CI** | | **p value** |
| Afghanistan | | Other | 32·3 | 15·7 | 48·9 | <0·001 | 27·5 | 14·2 | 40·7 | <0·001 |
| 2015 | | Turkmen | 29·8 | 16·4 | 43·2 |  | 26·2 | 15·4 | 37 |  |
|  | | Uzbek | 14·6 | 6·2 | 23·1 |  | 14·7 | 8·1 | 21·4 |  |
|  | | Baloch | 49 | 28·4 | 69·6 |  | 47·1 | 31·3 | 63 |  |
|  | | Hazara | 24·4 | 18·5 | 30·4 |  | 25·5 | 20·3 | 30·7 |  |
|  | | Nuristani | 98·5 | 96·7 | 100·3 |  | 89·9 | 83·7 | 96·1 |  |
|  | | Pashai | 37·4 | 23·4 | 51·3 |  | 34·4 | 21·6 | 47·3 |  |
|  | | Pashtun | 33·6 | 27·7 | 39·5 |  | 33·4 | 28·1 | 38·7 |  |
|  | | Tajik | 20·8 | 17·1 | 24·4 |  | 21·2 | 17·7 | 24·7 |  |
| Angola | | Other | 48·2 | 34·8 | 61·6 | <0·001 | 34·2 | 25·4 | 43 | <0·001 |
| 2015 | | Chokwe/kioko | 53·6 | 45 | 62·2 |  | 41 | 35 | 46·9 |  |
|  | | Kikongo/ukongo | 37·8 | 26·2 | 49·5 |  | 31·3 | 23·5 | 39 |  |
|  | | Kwanhama | 15·9 | 8·6 | 23·2 |  | 9·5 | 5·2 | 13·9 |  |
|  | | Muhumbi | 26·8 | 14·7 | 38·8 |  | 15·2 | 7·6 | 22·9 |  |
|  | | Nganguela | 66·2 | 46·9 | 85·6 |  | 42·7 | 30·5 | 54·9 |  |
|  | | Nhaneca | 42·5 | 29·5 | 55·6 |  | 23·7 | 16 | 31·3 |  |
|  | | Português | 21·1 | 17·9 | 24·4 |  | 30·1 | 26·2 | 34 |  |
|  | | Umbundu | 54·5 | 47·2 | 61·8 |  | 34·4 | 29·5 | 39·3 |  |
| Belize | | African descent | 12·7 | 4·6 | 20·7 | 0·069 | 16·4 | 6·3 | 26·4 | <0·001 |
| 2015 | | Indigenous | 11·3 | 0 | 24 |  | 8·5 | 1 | 15·9 |  |
|  | | Reference | 3·7 | 0·5 | 6·8 |  | 2·2 | 0·7 | 3·8 |  |
| Benin | | Other | 10·2 | 5·4 | 15 | <0·001 | 10·1 | 5·1 | 15·1 | <0·001 |
| 2017 | | Adja and related | 14·3 | 9·7 | 18·8 |  | 16·6 | 11·3 | 21·9 |  |
|  | | Bariba and related | 11 | 6·6 | 15·4 |  | 9·8 | 5·8 | 13·9 |  |
|  | | Betamaribe and related | 14·2 | 7·7 | 20·6 |  | 11·5 | 6·4 | 16·5 |  |
|  | | Dendi and related | 19·6 | 12·8 | 26·5 |  | 18·2 | 13·3 | 23·2 |  |
|  | | Fon and related | 8·1 | 5·9 | 10·3 |  | 9·4 | 6·9 | 12 |  |
|  | | Peulh and related | 50·9 | 41·5 | 60·2 |  | 34·3 | 27·5 | 41·1 |  |
|  | | Yoa, Lokpa and related | 13·3 | 4·8 | 21·8 |  | 14·7 | 5·4 | 24 |  |
|  | | Yoruba | 16·9 | 11·6 | 22·3 |  | 19·9 | 13·8 | 26 |  |
| Bosnia and Herzegovina | | Bosnian | 3·4 | 1·5 | 5·4 | 0·685 | 3·3 | 0 | 35·6 | 0·96 |
| 2011 | | Croat | 2·9 | 0 | 7·1 |  | 4·5 | 0 | 70 |  |
|  | | Serb | 2 | 0 | 4·3 |  | 2 | 0 | 15·1 |  |
| Burkina Faso | | Bissa | 0·9 | 0 | 2·7 | <0·001 | 1 | 0 | 3 | <0·001 |
| 2010 | | Bobo | 0·3 | 0 | 0·8 |  | 0·3 | 0 | 0·9 |  |
|  | | Dagara | 1·2 | 0 | 3·4 |  | 1·2 | 0 | 3·2 |  |
|  | | Fulfuldé / Peul | 8·4 | 2·7 | 14·1 |  | 7·4 | 2·5 | 12·3 |  |
|  | | Gourmatché | 13·1 | 6·7 | 19·5 |  | 11·6 | 5·7 | 17·6 |  |
|  | | Gourounsi | 0 | 0 | 0 |  | 0 | 0 | 0 |  |
|  | | Lobi | 5·3 | 0 | 13·2 |  | 5·2 | 0 | 12·9 |  |
|  | | Mossi | 3·5 | 2·1 | 4·8 |  | 3·5 | 2·1 | 4·9 |  |
|  | | Other | 3·7 | 0·1 | 7·4 |  | 4·1 | 0·1 | 8·1 |  |
|  | | Sénoufo | 16·4 | 7·1 | 25·8 |  | 17·2 | 7·7 | 26·7 |  |
|  | | Touareg / Bella | 31·8 | 14·2 | 49·4 |  | 28 | 11 | 45 |  |
| Central African Republic | | Banda | 44·9 | 37·2 | 52·6 | 0·111 | 44·3 | 37·3 | 51·3 | 0·001 |
| 2018 | | Gbaya | 48·9 | 43·5 | 54·2 |  | 46·5 | 41·6 | 51·4 |  |
|  | | Haoussa | 44·1 | 29·4 | 58·7 |  | 50·5 | 33·5 | 67·5 |  |
|  | | Mandja | 34·2 | 22·7 | 45·7 |  | 34·7 | 24·4 | 45 |  |
|  | | Mboum | 44·5 | 32·7 | 56·3 |  | 40·2 | 27·6 | 52·8 |  |
|  | | Ngbaka/Bantou | 39·9 | 29·5 | 50·4 |  | 48·9 | 37·6 | 60·2 |  |
|  | | Other | 39·6 | 28·5 | 50·7 |  | 42·8 | 31·3 | 54·3 |  |
|  | | Sara | 36·1 | 18·7 | 53·6 |  | 33·9 | 18·3 | 49·4 |  |
|  | | Yakoma/Sango | 58·2 | 45·8 | 70·6 |  | 65·9 | 55·1 | 76·8 |  |
| Cameroon | | Other | 18·1 | 14·6 | 21·5 | <0·001 | 16·9 | 13·9 | 19·9 | 0·6968 |
| 2018 | | Bamileke | 9 | 2·7 | 15·2 |  | 14·8 | 4·4 | 25·3 |  |
|  | | Bamoun | 9·8 | 3·6 | 15·9 |  | 12·2 | 4·6 | 19·8 |  |
|  | | Boulou | 9·4 | 2·6 | 16·2 |  | 15·6 | 4·3 | 26·9 |  |
|  | | Eton | 9·8 | 0·2 | 19·3 |  | 12·5 | 0 | 25·6 |  |
|  | | Ewondo | 8·4 | 3·3 | 13·5 |  | 12·1 | 4·6 | 19·7 |  |
|  | | Foulbe | 27·9 | 19·1 | 36·6 |  | 21·2 | 14·2 | 28·2 |  |
| Chad | | Arab | 50·3 | 41·4 | 59·2 | <0·001 | 51 | 41·9 | 60·1 | <0·001 |
| 2014 | | Other | 41·6 | 31·6 | 51·7 |  | 42·2 | 32·9 | 51·4 |  |
|  | | Sara | 26·5 | 21·7 | 31·3 |  | 27·5 | 22·6 | 32·4 |  |
|  | | Bidio/Migami/Kenga/Dangléat | 43·3 | 21·6 | 64·9 |  | 38·3 | 20·8 | 55·8 |  |
|  | | Boulala/Médégo/Kouka | 78·1 | 67·6 | 88·6 |  | 75·6 | 66·1 | 85·2 |  |
|  | | Dadajo/Kibet/Mouro | 40·4 | 24·5 | 56·3 |  | 39 | 22·7 | 55·3 |  |
|  | | Gorane | 54·6 | 42·7 | 66·5 |  | 54 | 40·8 | 67·2 |  |
|  | | Kanembou/Bornou/Boudouma | 57·7 | 47·5 | 68 |  | 54·4 | 44·8 | 64·1 |  |
|  | | Karo/Zimé/Pévé | 11·6 | 0·8 | 22·3 |  | 11·7 | 1·3 | 22·1 |  |
|  | | Marba/Lélé/Mesmé | 27·3 | 13 | 41·6 |  | 28·2 | 13·5 | 43 |  |
|  | | Massa/Mousseye/Mousgoume | 33·9 | 22·5 | 45·4 |  | 34·7 | 23 | 46·4 |  |
|  | | Moundang | 8·3 | 3·4 | 13·2 |  | 9·7 | 4·1 | 15·2 |  |
|  | | Ouadaï/Maba/Massalit/Mimi | 76·3 | 68·7 | 84 |  | 69·4 | 63·2 | 75·7 |  |
|  | | Peul/Foulbé/Bodoré | 57·7 | 41·6 | 73·9 |  | 54·6 | 42·2 | 67·1 |  |
|  | | Tama / Assongori / Mararit | 78 | 63·7 | 92·4 |  | 67·4 | 55·4 | 79·3 |  |
|  | | Toupouri/Kéra | 31·9 | 14·4 | 49·4 |  | 31·1 | 14·3 | 48 |  |
|  | | Zaghawa/Bideyat/Kobé | 78·6 | 65·5 | 91·7 |  | 80 | 61·2 | 98·8 |  |
| Colombia | | African descent | 3·2 | 1·3 | 5·2 | 0·472 | 3·2 | 1·2 | 5·1 | 0·498 |
| 2010 | | Indigenous | 4·4 | 1·5 | 7·3 |  | 4·5 | 1·1 | 7·8 |  |
|  | | Reference | 2·8 | 1·9 | 3·6 |  | 2·8 | 1·9 | 3·6 |  |
| Congo Brazzaville | | Kongo | 7·7 | 5·2 | 10·2 | <0·001 | 8·5 | 5·9 | 11·1 | <0·001 |
| 2014 | | Mbetis | 26·4 | 15·9 | 37 |  | 20·6 | 13·4 | 27·7 |  |
|  | | Mbosi | 15·6 | 8·6 | 22·5 |  | 16·1 | 9·7 | 22·5 |  |
|  | | Other | 18·5 | 13 | 24·1 |  | 17·3 | 12·3 | 22·3 |  |
|  | | Tekes | 25·7 | 18·6 | 32·9 |  | 22·5 | 17·1 | 27·9 |  |
|  | | Sangha-Likouala | 28·7 | 18·8 | 38·6 |  | 21·2 | 13·3 | 29·1 |  |
| Congo, DR | | Bantou | 34·1 | 29·9 | 38·4 | 0·134 | 34·7 | 30·5 | 38·8 | 0·144 |
| 2017 | | Nilotique | 19·9 | 7·6 | 32·1 |  | 16·8 | 4·7 | 29 |  |
|  | | Soudanais | 36·2 | 26·9 | 45·5 |  | 29·5 | 22·1 | 37 |  |
| Costa Rica | | African descent | 0·9 | 0·2 | 1·6 | 0·007 | 0·8 | 0·2 | 1·5 | 0·002 |
| 2018 | | Reference | 4·5 | 0·6 | 8·4 |  | 5·1 | 1 | 9·2 |  |
| Côte d'Ivoire | | Akan | 9·6 | 6·5 | 12·7 | <0·001 | 10·3 | 6·9 | 13·7 | 0·002 |
| 2016 | | Gur | 25·4 | 19·5 | 31·3 |  | 23·4 | 17·7 | 29·2 |  |
|  | | Krou | 17·2 | 7·2 | 27·2 |  | 19 | 7·7 | 30·3 |  |
|  | | Mandé du Nord | 23·8 | 16·8 | 30·9 |  | 26·4 | 18·8 | 34 |  |
|  | | Mandé du Sud | 21·7 | 13·1 | 30·3 |  | 19·6 | 11·9 | 27·2 |  |
|  | | Other | 23 | 18 | 28 |  | 21·5 | 16·7 | 26·4 |  |
| Cuba | | African descent | 5·1 | 0·3 | 10 | 0·025 | 4·5 | 0 | 9·2 | 0·0912 |
| 2019 | | Reference | 1·4 | 0·5 | 2·3 |  | 1·5 | 0·4 | 2·5 |  |
| Dominican Republic | | African descent | 21·7 | 16·1 | 27·3 | <0·001 | 14·9 | 10·7 | 19·2 | <0·001 |
| 2014 | | Reference | 7·6 | 6·3 | 8·9 |  | 7·9 | 6·6 | 9·3 |  |
| Ethiopia | | Other | 25·5 | 17·1 | 34 | <0·001 | 26·2 | 17·5 | 35 | <0·001 |
| 2016 | | Affar | 51·5 | 31·9 | 71·2 |  | 43·2 | 29·3 | 57·1 |  |
|  | | Amhara | 18·8 | 12·5 | 25·1 |  | 20·6 | 13·9 | 27·2 |  |
|  | | Berta | 6·1 | 1·5 | 10·6 |  | 5·7 | 1·6 | 9·9 |  |
|  | | Guragie | 13·3 | 0 | 28·4 |  | 17·5 | 0·3 | 34·6 |  |
|  | | Nuwer | 55·4 | 28·9 | 81·8 |  | 51·3 | 27·6 | 75·1 |  |
|  | | Oromo | 34·3 | 27·2 | 41·3 |  | 32·5 | 25·8 | 39·2 |  |
|  | | Somalie | 36·7 | 27·7 | 45·7 |  | 30·6 | 21·9 | 39·3 |  |
|  | | Tigrie | 7·7 | 2·1 | 13·4 |  | 8·2 | 2·4 | 14 |  |
| Gabon | | Other | 11·8 | 5·3 | 18·3 | 0·849 | 11·1 | 5·4 | 16·8 | 0·626 |
| 2012 | | Fang | 15·1 | 8·7 | 21·5 |  | 18·2 | 10 | 26·5 |  |
|  | | Kota-Kele | 13·7 | 3·7 | 23·6 |  | 12·1 | 4·1 | 20·2 |  |
|  | | Mbede-Teke | 12 | 2·3 | 21·7 |  | 9·8 | 1·6 | 18·1 |  |
|  | | Nzabi-Duma | 8·9 | 3·2 | 14·6 |  | 9·1 | 3·4 | 14·8 |  |
|  | | Shira-Punu/Vili | 10 | 1 | 19 |  | 10 | 0·6 | 19·5 |  |
| Gambia | | Fula | 5·5 | 1·6 | 9·3 | 0·07 | 5·7 | 1·6 | 9·8 | 0·069 |
| 2018 | | Jola | 2·1 | 0 | 6·3 |  | 1·8 | 0 | 5·3 |  |
|  | | Mandinka | 1·2 | 0·1 | 2·2 |  | 1·2 | 0·1 | 2·3 |  |
|  | | Other | 5·8 | 2·3 | 9·4 |  | 5·4 | 1·9 | 8·8 |  |
|  | | Sarahule | 2·6 | 0·2 | 5 |  | 2·5 | 0·1 | 5 |  |
|  | | Wollof | 2·7 | 0 | 6·1 |  | 2·9 | 0 | 6·7 |  |
| Ghana | | Akan | 3·9 | 1·5 | 6·3 | 0·577 | 4·4 | 1·6 | 7·1 | 0·329 |
| 2017 | | Ewe | 2·1 | 0 | 4·5 |  | 2·2 | 0 | 4·6 |  |
|  | | Gruma | 5·7 | 0·2 | 11·2 |  | 4·2 | 0 | 8·6 |  |
|  | | Grusi | 5 | 0 | 11·6 |  | 4 | 0 | 9·2 |  |
|  | | Guan | 2·4 | 0 | 7·2 |  | 2 | 0 | 6·2 |  |
|  | | Mole Dagbani | 3 | 0 | 6 |  | 2·6 | 0 | 5·2 |  |
|  | | Other | 6·3 | 1·9 | 10·8 |  | 6 | 1·8 | 10·3 |  |
|  | | Ga/Damgme | 7 | 1·3 | 12·8 |  | 8·5 | 1·6 | 15·5 |  |
| Guatemala | | Indigenous | 3·8 | 2·5 | 5·1 | 0·003 | 3·7 | 2·2 | 5·3 | 0·012 |
| 2014 | | Reference | 1·3 | 0·5 | 2·1 |  | 1·3 | 0·5 | 2·2 |  |
| Guinea | | Guerzé | 15·3 | 5·6 | 25 | <0·001 | 14·7 | 5·6 | 23·9 | <0·001 |
| 2018 | | Kissi | 32·9 | 23·1 | 42·7 |  | 31·5 | 22·4 | 40·5 |  |
|  | | Malinké | 30·5 | 24·2 | 36·8 |  | 31·6 | 25 | 38·3 |  |
|  | | Peulh | 53·8 | 48·2 | 59·4 |  | 49·9 | 44·9 | 54·9 |  |
|  | | Soussou | 28·7 | 22·6 | 34·8 |  | 32·1 | 25·9 | 38·3 |  |
| Guinea Bissau | | Balanta | 8·3 | 3·9 | 12·7 | 0·166 | 8·6 | 3·8 | 13·4 | 0·22 |
| 2018 | | Beafada | 4·4 | 0 | 9·8 |  | 5·7 | 0 | 12·9 |  |
|  | | Fula | 8 | 3·9 | 12·2 |  | 7·6 | 3·5 | 11·7 |  |
|  | | Mandinga | 5·6 | 2·3 | 8·8 |  | 5·4 | 2·5 | 8·3 |  |
|  | | Manjaco | 5·1 | 0·5 | 9·8 |  | 5·9 | 0·5 | 11·3 |  |
|  | | Other | 10·6 | 0·5 | 20·7 |  | 10·7 | 0·9 | 20·5 |  |
|  | | Papel | 1·1 | 0 | 2·7 |  | 1·2 | 0 | 2·9 |  |
| Guyana | | African descent | 4·5 | 1·2 | 7·8 | 0·658 | 4·4 | 1·4 | 7·5 | 0·947 |
| 2014 | | Indigenous | 6·2 | 2·3 | 10·1 |  | 4·9 | 0·8 | 9 |  |
|  | | Reference | 4 | 0·8 | 7·1 |  | 4·2 | 1 | 7·3 |  |
| Honduras | | African descent | 0·8 | 0 | 2·5 | 0·106 | 0·9 | 0 | 2·6 | 0·073 |
| 2011 | | Indigenous | 0·1 | 0 | 0·3 |  | 0·1 | 0 | 0·4 |  |
|  | | Reference | 1 | 0·5 | 1·5 |  | 1 | 0·5 | 1·5 |  |
| India | | Caste | 10·2 | 9·8 | 10·7 | <0·001 | 10·4 | 9·9 | 10·8 | 0·045 |
| 2015 | | No caste/Tribe | 11 | 8·6 | 13·4 |  | 11·7 | 9·1 | 14·2 |  |
|  | | Tribe | 14·2 | 12·6 | 15·8 |  | 12 | 10·6 | 13·4 |  |
| Iraq | | Arabic | 14·5 | 12·2 | 16·7 | 0·015 | 13·7 | 11·7 | 15·8 | 0·195 |
| 2018 | | Kurdish | 6·7 | 2·7 | 10·7 |  | 9·1 | 3·7 | 14·5 |  |
| Jordan | | Other | 19·5 | 3 | 36·1 | 0·009 | 13·5 | 4·6 | 22·3 | 0·137 |
| 2017 | | Jordanian | 6·4 | 4·5 | 8·3 |  | 6·9 | 4·9 | 8·9 |  |
|  | | Syrian | 10·7 | 4·4 | 17·1 |  | 7·9 | 2·5 | 13·4 |  |
| Kazakhstan | | Kazakh | 3 | 1·8 | 4·3 | 0·001 | 3 | 1·8 | 4·2 | 0·001 |
| 2015 | | Other | 4·3 | 0·5 | 8·1 |  | 5·3 | 0·4 | 10·2 |  |
|  | | Russian | 12·8 | 5·1 | 20·6 |  | 10·8 | 4·9 | 16·7 |  |
| Kenya | | Other | 1·1 | 0 | 2·4 | <0·001 | 0·9 | 0 | 2 | 0·284 |
| 2014 | | Boran | 1·8 | 0 | 4·3 |  | 1·2 | 0 | 2·8 |  |
|  | | Kalenjin | 2·8 | 1·4 | 4·2 |  | 3 | 1·4 | 4·6 |  |
|  | | Kamba | 0·9 | 0 | 2·1 |  | 1·2 | 0 | 2·7 |  |
|  | | Kikuya | 2·1 | 0 | 4·5 |  | 3·1 | 0 | 6·2 |  |
|  | | Kisii | 0·6 | 0 | 1·6 |  | 1 | 0 | 2·5 |  |
|  | | Luhya | 2·1 | 0 | 4·5 |  | 3 | 0 | 6·3 |  |
|  | | Luo | 1·7 | 0·2 | 3·2 |  | 2·3 | 0·1 | 4·4 |  |
|  | | Maasai | 6 | 1·7 | 10·4 |  | 2·7 | 0·6 | 4·8 |  |
|  | | Meru | 0·4 | 0 | 1·1 |  | 0·5 | 0 | 1·6 |  |
|  | | Mijikenda/Swahili | 3·6 | 0 | 7·5 |  | 2·5 | 0 | 5·4 |  |
|  | | Pokomo | 1·3 | 0 | 3·9 |  | 1·1 | 0 | 3·5 |  |
|  | | Samburu | 6·3 | 0 | 13·2 |  | 2·4 | 0 | 5·2 |  |
|  | | Somali | 11 | 6·6 | 15·5 |  | 4 | 1·5 | 6·6 |  |
|  | | Turkana | 4·8 | 0·9 | 8·6 |  | 1·7 | 0 | 3·3 |  |
| Kyrgyzstan | | Kyrgyz | 8 | 4·8 | 11·2 | 0·179 | 7·9 | 4·7 | 11 | 0·132 |
| 2018 | | Uzbek | 13·5 | 4·8 | 22·1 |  | 15·1 | 4·7 | 25·6 |  |
| Laos | | Chinese-Tibetan | 36·3 | 23·7 | 48·9 | <0·001 | 28·2 | 18·4 | 37·9 | <0·001 |
| 2017 | | Hmong-Mien | 42·9 | 36·5 | 49·3 |  | 36·5 | 31·1 | 41·9 |  |
|  | | Lao-Tai | 19 | 16·1 | 21·9 |  | 22·5 | 19·1 | 26 |  |
|  | | Mon-Khmer | 34·4 | 28·9 | 40 |  | 28·3 | 23·9 | 32·7 |  |
| Malawi | | Other | 4·9 | 0·4 | 9·4 | 0·136 | 5·2 | 0·3 | 10·1 | 0·077 |
| 2015 | | Chewa | 1·6 | 0·2 | 2·9 |  | 1·5 | 0·3 | 2·8 |  |
|  | | Lomwe | 3·9 | 1·6 | 6·2 |  | 4·1 | 1·7 | 6·6 |  |
|  | | Mang'Anja | 2·4 | 0 | 5·7 |  | 2·3 | 0 | 5·5 |  |
|  | | Ngoni | 2·3 | 0·1 | 4·6 |  | 2·4 | 0·1 | 4·8 |  |
|  | | Sena | 7·1 | 1·9 | 12·3 |  | 6·3 | 1·8 | 10·7 |  |
|  | | Tonga | 0·9 | 0 | 2·8 |  | 1·1 | 0 | 3·2 |  |
|  | | Tumbuka | 3·2 | 0·8 | 5·6 |  | 3·9 | 0·9 | 7 |  |
|  | | Yao | 1·9 | 0·7 | 3·1 |  | 1·7 | 0·6 | 2·8 |  |
| Mali | | Other | 11·8 | 5·8 | 17·9 | <0·001 | 12 | 5·8 | 18·2 | <0·001 |
| 2018 | | Bambara | 15·3 | 10·5 | 20·1 |  | 15·9 | 11·1 | 20·7 |  |
|  | | Dogon | 20·9 | 8·7 | 33 |  | 17·4 | 7·2 | 27·6 |  |
|  | | Malinke | 27·2 | 16·2 | 38·2 |  | 29·2 | 18 | 40·4 |  |
|  | | Peulh | 22 | 14·9 | 29·2 |  | 21·9 | 15·3 | 28·5 |  |
|  | | Sarakole/Soninke/Marka | 8·2 | 3·5 | 12·9 |  | 8·4 | 3·5 | 13·4 |  |
|  | | Sonraï | 29·8 | 20·3 | 39·2 |  | 31·5 | 21·6 | 41·4 |  |
|  | | Sénoufo/Minianka | 10·4 | 4·3 | 16·5 |  | 10·3 | 4·3 | 16·3 |  |
|  | | Touareg/Bella | 51·7 | 32·1 | 71·3 |  | 41·9 | 29 | 54·8 |  |
| Mauritania | | Arabe | 14·7 | 12·1 | 17·4 | 0·06 | 14·5 | 12·1 | 16·9 | 0·013 |
| 2015 | | Poular | 10·8 | 6·9 | 14·7 |  | 11 | 7·1 | 14·9 |  |
|  | | Soninké | 24·8 | 8·6 | 41 |  | 34 | 9·5 | 58·4 |  |
| Mexico | | Indigenous | 10·5 | 3·9 | 17·1 | 0·365 | 9·6 | 3·5 | 15·8 | 0·535 |
| 2015 | | Reference | 7·5 | 4·8 | 10·2 |  | 7·6 | 4·9 | 10·3 |  |
| Mongolia | | Kazakh | 12 | 3·7 | 20·2 | 0·001 | 11·7 | 3·2 | 20·2 | <0·001 |
| 2018 | | Khalkh | 2·6 | 1·2 | 4 |  | 2·6 | 1·2 | 4 |  |
|  | | Other | 1·8 | 0 | 4·4 |  | 1·9 | 0 | 4·5 |  |
| Montenegro | | Montenegrin | 3·5 | 0·3 | 6·7 | 0·338 | 3·7 | 0·2 | 7·3 | 0·448 |
| 2013 | | Other | 10·5 | 0 | 22·3 |  | 9·4 | 0·1 | 18·7 |  |
|  | | Serbian | 6·5 | 0 | 15·3 |  | 6·8 | 0 | 14·9 |  |
| Mozambique | | Other | 11·2 | 6·1 | 16·2 | 0·001 | 10·9 | 6 | 15·9 | 0·168 |
| 2015 | | Cindau | 10·8 | 1·6 | 20 |  | 11·1 | 1·9 | 20·2 |  |
|  | | Cinyanja | 26·8 | 9 | 44·5 |  | 20·7 | 6·1 | 35·2 |  |
|  | | Cisena | 11·8 | 4·4 | 19·2 |  | 12·5 | 5·3 | 19·8 |  |
|  | | Emakhuwa | 10·7 | 0·8 | 20·5 |  | 9·1 | 0·9 | 17·3 |  |
|  | | Português | 1·1 | 0 | 3·4 |  | 1·9 | 0 | 5·8 |  |
|  | | Xichangana | 1·4 | 0 | 3 |  | 2·4 | 0 | 5·5 |  |
|  | | Xitswa | 3·8 | 0 | 10·7 |  | 4·6 | 0 | 12·6 |  |
| Myanmar | | Other | 26 | 15·4 | 36·7 | <0·001 | 18·1 | 11·2 | 25 | 0·074 |
| 2015 | | Myanmar | 10·7 | 7·7 | 13·7 |  | 11·7 | 8·3 | 15 |  |
| Namibia | | Other | 8·2 | 0 | 17·2 | 0·416 | 8 | 0 | 16·6 | 0·506 |
| 2013 | | Afrikaans | 6·4 | 0 | 14·6 |  | 4·1 | 0 | 9·4 |  |
|  | | Damara/Nama | 6·2 | 0·6 | 11·9 |  | 4·9 | 0·6 | 9·3 |  |
|  | | Herero | 18·9 | 3 | 34·7 |  | 15 | 2·3 | 27·7 |  |
|  | | Kwangali | 8·3 | 2 | 14·7 |  | 10·2 | 2·2 | 18·1 |  |
|  | | Lozi | 6·5 | 0 | 13·4 |  | 7·7 | 0·2 | 15·3 |  |
|  | | Oshiwambo | 5·6 | 2·9 | 8·3 |  | 6·3 | 3·3 | 9·2 |  |
| Nepal | | Brahman - Hill | 9·3 | 3·7 | 14·8 | 0·049 | 14·2 | 6·4 | 22 | 0·102 |
| 2019 | | Chhetree | 11·4 | 6·8 | 16 |  | 11·5 | 6·9 | 16·1 |  |
|  | | Kami | 6·9 | 1·3 | 12·5 |  | 6·6 | 1·3 | 11·9 |  |
|  | | Magar | 5·1 | 1·2 | 9·1 |  | 6·9 | 1·3 | 12·5 |  |
|  | | Musalman | 21·2 | 8·6 | 33·7 |  | 17·5 | 6·5 | 28·4 |  |
|  | | Other | 12 | 8·2 | 15·9 |  | 10·9 | 7·6 | 14·2 |  |
|  | | Tamang | 4·3 | 0 | 9·2 |  | 4·2 | 0 | 8·9 |  |
|  | | Tharu | 5 | 0 | 10·8 |  | 5·1 | 0 | 10·5 |  |
| Niger | | Haoussa | 15·1 | 12 | 18·3 | <0·001 | 14·8 | 11·8 | 17·9 | <0·001 |
| 2012 | | Kanouri/Toubou | 39·5 | 29·3 | 49·7 |  | 35·6 | 27·1 | 44·2 |  |
|  | | Other | 25·5 | 15·2 | 35·8 |  | 24·8 | 15·2 | 34·5 |  |
|  | | Zarma | 5 | 2·6 | 7·5 |  | 5·5 | 2·8 | 8·2 |  |
| Nigeria | | Other | 24·3 | 21·1 | 27·5 | <0·001 | 25·9 | 22·7 | 29 | <0·001 |
| 2018 | | Fulani | 63·5 | 58 | 69 |  | 43·4 | 39·5 | 47·4 |  |
|  | | Hausa | 55·1 | 51·6 | 58·7 |  | 41·7 | 38·8 | 44·5 |  |
|  | | Ibibio | 15·5 | 8·1 | 22·9 |  | 25·7 | 13·7 | 37·8 |  |
|  | | Igala | 33·9 | 18·3 | 49·5 |  | 45·4 | 26·6 | 64·2 |  |
|  | | Igbo | 6·3 | 4·3 | 8·4 |  | 12 | 8·1 | 15·9 |  |
|  | | Ijaw/Izon | 20·7 | 12·3 | 29·2 |  | 36·1 | 23·4 | 48·7 |  |
|  | | Kanuri/Beriberi | 51·9 | 42·4 | 61·5 |  | 40·3 | 31·6 | 49 |  |
|  | | Tiv | 27·8 | 17·9 | 37·8 |  | 30 | 21 | 38·9 |  |
|  | | Yoruba | 12·7 | 8·9 | 16·6 |  | 23·9 | 16·9 | 30·9 |  |
| North Macedonia | | Albanian | 2·9 | 0 | 8·6 | 0·567 | 3·5 | 0 | 10·3 | 0·437 |
| 2018 | | Macedonian | 1·6 | 0 | 3·2 |  | 1·5 | 0 | 3 |  |
| Pakistan | | Other | 17·2 | 11 | 23·5 | <0·001 | 15·9 | 9·6 | 22·1 | <0·001 |
| 2017 | | Baluchi | 27·9 | 16·6 | 39·3 |  | 20·2 | 12·9 | 27·5 |  |
|  | | Punjabi | 2·9 | 0·9 | 5 |  | 4·3 | 1·3 | 7·3 |  |
|  | | Pushto | 28·3 | 21·3 | 35·3 |  | 23·9 | 18·5 | 29·3 |  |
|  | | Sariaki | 15·3 | 7·2 | 23·3 |  | 11·3 | 5·8 | 16·8 |  |
|  | | Sindhi | 26·7 | 14·9 | 38·5 |  | 18·1 | 10·2 | 26 |  |
|  | | Urdu | 0·9 | 0 | 2·2 |  | 1·8 | 0 | 4·6 |  |
| Panama | | African descent | 4·9 | 1 | 8·8 | 0·02 | 3·9 | 0·8 | 7 | 0·004 |
| 2013 | | Indigenous | 13·9 | 8·4 | 19·3 |  | 17·2 | 7·9 | 26·5 |  |
|  | | Reference | 6·4 | 2·7 | 10 |  | 6·3 | 2·8 | 9·8 |  |
| Papua New Guinea | | English | 14·7 | 0·7 | 28·7 | 0·148 | 27·1 | 0 | 57·4 | 0·021 |
| 2016 | | Pidgin | 38·3 | 31·9 | 44·6 |  | 42·2 | 36·6 | 47·8 |  |
|  | | Tok Ples | 36·1 | 31·2 | 41·1 |  | 33·3 | 29·3 | 37·4 |  |
| Paraguay | | Indigenous | 6·8 | 4·1 | 9·4 | 0·085 | 6·6 | 3·7 | 9·4 | 0·161 |
| 2016 | | Reference | 4·1 | 2·4 | 5·7 |  | 4·2 | 2·5 | 5·9 |  |
| Peru | | Indigenous | 6·7 | 3·5 | 9·8 | 0·156 | 5 | 2·4 | 7·7 | 0·818 |
| 2019 | | Reference | 4·6 | 3·7 | 5·5 |  | 4·7 | 3·6 | 5·8 |  |
| Philippines | | Other | 18·9 | 14·6 | 23·2 | <0·001 | 15·5 | 12 | 19·1 | <0·001 |
| 2017 | | Bikolano | 13·9 | 4·8 | 22·9 |  | 14·3 | 4·9 | 23·7 |  |
|  | | Cebuano | 13·4 | 8·5 | 18·4 |  | 13·5 | 8·8 | 18·2 |  |
|  | | Ilokano | 9·5 | 4·2 | 14·7 |  | 10·2 | 4·3 | 16·1 |  |
|  | | Ilonggo | 15·1 | 7·7 | 22·6 |  | 16·7 | 8·3 | 25 |  |
|  | | Maranao | 63·9 | 49·1 | 78·6 |  | 40·5 | 30 | 51 |  |
|  | | Tagalog | 7·9 | 4·1 | 11·6 |  | 9·5 | 5·1 | 13·9 |  |
|  | | Visaya | 8·7 | 1·8 | 15·7 |  | 8·3 | 1·6 | 15 |  |
|  | | Waray | 3·9 | 0·5 | 7·3 |  | 3·6 | 0·4 | 6·7 |  |
| Senegal | | Other | 5·7 | 0 | 13·2 | 0·272 | 8·3 | 0 | 17·6 | 0·3482 |
| 2019 | | Poular | 5·7 | 2·3 | 9·2 |  | 4·4 | 2 | 6·7 |  |
|  | | Wolof | 2·7 | 1·1 | 4·2 |  | 3 | 1·4 | 4·6 |  |
|  | | Mandingue/ Socé | 2·5 | 0 | 5·6 |  | 3·2 | 0 | 7 |  |
|  | | Serer | 2·1 | 0 | 4·2 |  | 2 | 0·1 | 3·9 |  |
| Sierra Leone | | Other | 2·4 | 0·7 | 4·1 | <0·001 | 2·4 | 0·7 | 4·1 | <0·001 |
| 2019 | | Fullah | 0 | 0 | 0 |  | 0 | 0 | 0 |  |
|  | | Kono | 0 | 0 | 0 |  | 0 | 0 | 0 |  |
|  | | Korankoh | 8·9 | 0·3 | 17·5 |  | 8·5 | 0·3 | 16·7 |  |
|  | | Limba | 6·5 | 1 | 11·9 |  | 6·7 | 1·3 | 12·2 |  |
|  | | Mende | 6 | 3·6 | 8·5 |  | 5·9 | 3·5 | 8·2 |  |
|  | | Temne | 6 | 3·8 | 8·2 |  | 6·1 | 3·9 | 8·4 |  |
| South Africa | | Black/African | 9 | 5·9 | 12·2 | 0·722 | 9 | 5·8 | 12·2 | 0·826 |
| 2016 | | Coloured | 7·5 | 0·5 | 14·6 |  | 7·9 | 0·2 | 15·7 |  |
| Suriname | | African descent | 22·6 | 15·3 | 29·9 | 0·35 | 20·9 | 12·6 | 29·3 | 0·699 |
| 2018 | | Reference | 18·3 | 12·4 | 24·2 |  | 18·9 | 13 | 24·8 |  |
| Tajikistan | | Other | 6·3 | 2·2 | 10·5 | 0·524 | 6·8 | 2·4 | 11·2 | 0·7 |
| 2017 | | Russian | 7·9 | 6·2 | 9·5 |  | 7·7 | 6·1 | 9·4 |  |
| Thailand | | Non-Thai | 15·2 | 1·6 | 28·8 | <0·001 | 17·9 | 5·8 | 30 | <0·001 |
| 2019 | | Thai | 2 | 0·8 | 3·2 |  | 2 | 0·8 | 3·1 |  |
| Timor Leste | | Other | 15·2 | 4·4 | 25·9 | 0·307 | 12·4 | 4·3 | 20·5 | 0·081 |
| 2016 | | Tetum | 22 | 19 | 25 |  | 22·3 | 19·6 | 25·1 |  |
| Togo | | Adja-Ewe | 10·3 | 5·8 | 14·8 | 0·408 | 12·7 | 7·8 | 17·7 | 0·014 |
| 2017 | | Akposso/Akébou | 8·2 | 5·1 | 11·4 |  | 5·5 | 3·3 | 7·8 |  |
|  | | Kabye-Tem | 5·5 | 1·5 | 9·6 |  | 6·5 | 1·9 | 11·1 |  |
|  | | Other | 12·4 | 3·7 | 21·2 |  | 13·3 | 5·1 | 21·6 |  |
| Turkmenistan | | Turkmen | 0·8 | 0·1 | 1·4 | <0·001 | 0·7 | 0·1 | 1·3 | <0·001 |
| 2015 | | Uzbek | 0 | 0 | 0 |  | 0 | 0 | 0 |  |
| Uganda | | Other | 4·6 | 2·1 | 7·1 | 0·072 | 4·2 | 1·8 | 6·6 | 0·027 |
| 2016 | | Acholi | 0·9 | 0 | 2·3 |  | 0·9 | 0 | 2·2 |  |
|  | | Alur | 4·5 | 0·6 | 8·3 |  | 3·9 | 0·5 | 7·2 |  |
|  | | Bafumbira | 0·5 | 0 | 1·6 |  | 0·5 | 0 | 1·4 |  |
|  | | Baganda | 6 | 2·7 | 9·2 |  | 6·5 | 2·8 | 10·2 |  |
|  | | Bagisu | 3·8 | 0·3 | 7·3 |  | 4·1 | 0·4 | 7·7 |  |
|  | | Bagwere | 11 | 1·5 | 20·5 |  | 11·5 | 1·8 | 21·2 |  |
|  | | Bakiga | 7·6 | 3·3 | 11·8 |  | 7·5 | 3·1 | 11·9 |  |
|  | | Bakonzo | 3·2 | 0 | 7·3 |  | 3·3 | 0 | 7·6 |  |
|  | | Banyankore | 4·1 | 1·1 | 7·1 |  | 4·1 | 1·1 | 7·1 |  |
|  | | Banyarwanda | 3·2 | 0 | 8 |  | 2·8 | 0 | 7 |  |
|  | | Banyole | 15·2 | 0·5 | 29·9 |  | 15·2 | 0·9 | 29·5 |  |
|  | | Banyoro | 4·4 | 0 | 8·9 |  | 4·8 | 0 | 9·8 |  |
|  | | Basoga | 7·4 | 3·2 | 11·7 |  | 7·9 | 3·3 | 12·6 |  |
|  | | Batoro | 8·6 | 2 | 15·2 |  | 8·9 | 2 | 15·8 |  |
|  | | Iteso | 3·4 | 1·2 | 5·6 |  | 3·6 | 1·3 | 5·9 |  |
|  | | Jopadhola | 1·9 | 0 | 4·7 |  | 1·9 | 0 | 4·8 |  |
|  | | Karimojong | 2·3 | 0 | 5·2 |  | 1·5 | 0 | 3·3 |  |
|  | | Lango | 5 | 0 | 10 |  | 5 | 0·1 | 10 |  |
|  | | Lugbara | 5 | 0 | 10·7 |  | 4·8 | 0 | 10·1 |  |
| Vietnam | | Kinh | 2·4 | 1 | 3·7 | <0·001 | 3·4 | 1·5 | 5·3 | 0·589 |
| 2013 | | Non-Kinh | 11·9 | 6·1 | 17·7 |  | 4·3 | 1·9 | 6·6 |  |
| Zambia | | Other | 3·1 | 0·3 | 6 | 0·099 | 3 | 0·6 | 5·4 | 0·146 |
| 2018 | | Bemba | 1·9 | 0·5 | 3·4 |  | 1·9 | 0·6 | 3·2 |  |
|  | | Kaonde | 0·6 | 0 | 1·8 |  | 0·7 | 0 | 2·1 |  |
|  | | Lozi | 3·7 | 1·1 | 6·2 |  | 3·5 | 1·1 | 5·9 |  |
|  | | Lunda | 10·3 | 0 | 22·7 |  | 11·8 | 0 | 27·2 |  |
|  | | Luvale | 1·9 | 0 | 5·6 |  | 2·6 | 0 | 7·6 |  |
|  | | Nyanja | 1·8 | 0·4 | 3·2 |  | 1·7 | 0·4 | 3 |  |
|  | | Tonga | 0·9 | 0 | 2·1 |  | 1 | 0 | 2·3 |  |
| Zimbabwe | | Other | 0 | 0 | 0 | <0·001 | 0 | 0 | 0 | <0·001 |
| 2019 | | Ndebele | 1·8 | 0·1 | 3·6 |  | 1·7 | 0 | 3·4 |  |
|  | | Shona | 6·3 | 4·2 | 8·4 |  | 6·5 | 4·3 | 8·6 |  |

Legend: no-DPT: children who did not received any doses of the diphtheria-tetanus-pertussis-containing vaccine. CI: confidence interval.

* Adjustment variables: household wealth quintiles, maternal education, and area of residence.

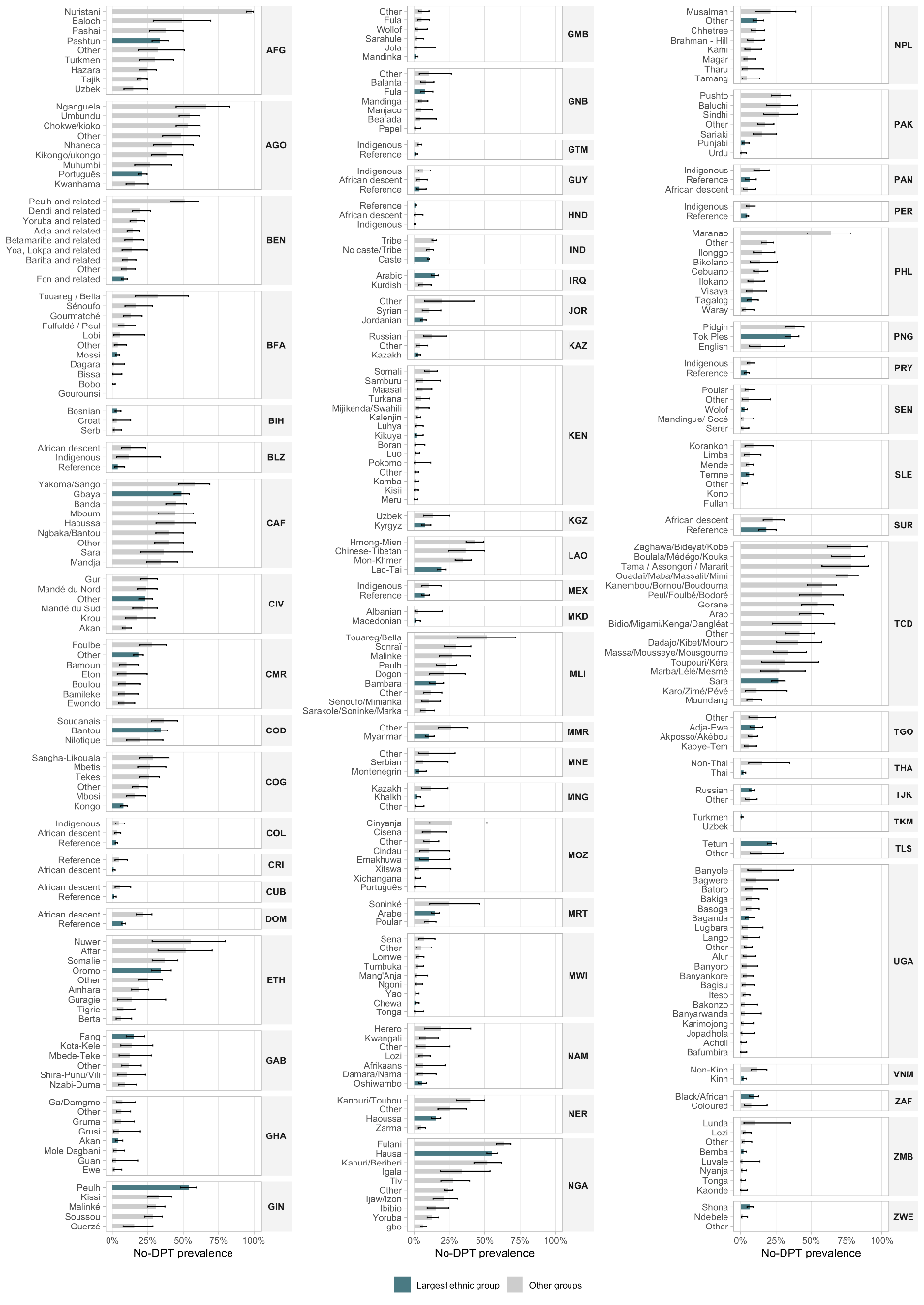

Supplementary Figure 1. No-DPT prevalence according to ethnic group.

Legend: AFG: Afghanistan, AGO: Angola, BLZ: Belize, BEN: Benin, BIH: Bosnia and Herzegovina, BFA: Burkina Faso, CAF: Central African Republic, CMR: Cameroon, TCD: Chad,	COL: Colombia, COG: Congo Brazzaville, COD: Democratic Republic of Congo, CRI: Costa Rica, CIV: Cotê d'Ivoire, CUB: Cuba, DOM: Dominican Republic, ETH: Ethiopia, GAB: Gabon, GMB: Gambia, GHA: Ghana, GTM: Guatemala, GIN: Guinea,	GNB: Guinea Bissau, GUY: Guyana, HND: Honduras, IND: India, IRQ: Iraq, JOR: Jordan, KAZ: Kazakhstan, KEN: Kenya, KGZ: Kyrgyzstan, LAO: Laos, MWI: Malawi, MLI: Mali, MRT: Mauritania, MEX: Mexico, MNG: Mongolia, MNE: Montenegro, MOZ: Mozambique, MMR: Myanmar, NAM: Namibia, NPL: Nepal, NER: Niger, NGA: Nigeria, MKD: North Macedonia, PAK: Pakistan, PAN: Panama, PNG: Papua New Guinea, PRY: Paraguay, PER: Peru, PHL: Philippines, SEN: Senegal, SLE: Sierra Leone, ZAF: South Africa, SUR: Suriname, TJK: Tajikistan, THA: Thailand, TLS: Timor-Leste, TGO: Togo, TKM: Turkmenistan, UGA: Uganda, VNM: Vietnam, ZMB: Zambia, ZWE: Zimbabwe.

### Supplementary Table 4 – No-DPT prevalence in ethnic groups present in more than one country (limited to groups with prevalence >10% in at least one country).

| **Country** | **Year** | **Ethnic group** | **No-DPT** | **National no-DPT** | **Difference %ethnic - %national (pp)** |
| --- | --- | --- | --- | --- | --- |
| Afghanistan | 2015 | Baloch/Baluchi | 49·0% | 27·0% | 22·0 |
| Pakistan | 2017 |  | 27·9% | 13·7% | 14·2 |
| Burkina Faso | 2010 | Sénoufo/Minianka | 16·3% | 5·6% | 10·7 |
| Mali | 2018 |  | 10·4% | 17·9% | -7·5 |
| Burkina Faso | 2010 | Touareg / Bella | 31·8% | 5·6% | 26·2 |
| Mali | 2018 |  | 51·7% | 17·9% | 33·8 |
| Central African Republic | 2018 | Haoussa/Hausa | 44·1% | 45·0% | -0·9 |
| Niger | 2012 |  | 15·1% | 13·8% | 1·3 |
| Nigeria | 2018 |  | 55·1% | 34·7% | 20·4 |
| Chad | 2014 | Arab/Arabic/Arabe | 50·3% | 41·7% | 8·6 |
| Iraq | 2018 |  | 14·5% | 13·3% | 1·2 |
| Mauritania | 2015 |  | 14·7% | 14·2% | 0·5 |
| Ethiopia | 2016 | Somalie/somali | 36·7% | 26·8% | 9·9 |
| Kenya | 2014 |  | 11·0% | 2·5% | 8·5 |
| Benin | 2017 | Fula/Fulani/Fullah/Peulh/Fulfuldé / Peul/ Foulbe | 50·9% | 15·8% | 35·1 |
| Burkina Faso | 2010 |  | 8·4% | 5·6% | 2·8 |
| Cameroon | 2018 |  | 27·9% | 16·7% | 11·2 |
| Chad | 2014 |  | 57·7% | 41·7% | 16·0 |
| Gambia | 2018 |  | 5·5% | 3·2% | 2·3 |
| Guinea | 2018 |  | 53·8% | 37·7% | 16·1 |
| Guinea Bissau | 2018 |  | 8·0% | 7·0% | 1·0 |
| Mali | 2018 |  | 22·0% | 17·9% | 4·1 |
| Nigeria | 2018 |  | 63·5% | 34·7% | 28·8 |
| Sierra Leone | 2019 |  | 0·0% | 5·4% | -5·4 |
| Gambia | 2018 | Mandinka/Mandinga/Malinke | 1·2% | 3·2% | -2·0 |
| Guinea | 2018 |  | 30·5% | 37·7% | -7·2 |
| Guinea Bissau | 2018 |  | 5·6% | 7·0% | -1·4 |
| Mali | 2018 |  | 27·2% | 17·9% | 9·3 |
| Benin | 2017 | Adja/Ewe | 14·3% | 15·8% | -1·5 |
| Togo | 2017 |  | 10·3% | 9·2% | 1·1 |
| Benin | 2017 | Yoruba | 16·9% | 15·8% | 1·1 |
| Nigeria | 2018 |  | 12·7% | 34·7% | -22·0 |
| Gambia | 2018 | Sarahule/Sarakole/Soninke/Marka | 2·6% | 3·2% | -0·6 |
| Mauritania | 2015 |  | 24·8% | 14·2% | 10·6 |
| Mali | 2018 |  | 8·2% | 17·9% | -9·7 |
| Niger | 2012 | Kanouri/Toubou/Kanuri/Beriberi | 39·5% | 13·8% | 25·7 |
| Nigeria | 2018 |  | 51·9% | 34·7% | 17·2 |

Legend: No-DPT: children who did not received any doses of the diphtheria-tetanus-pertussis-containing vaccine. pp: percentage points.

### Supplementary Table 5 – Proportion of children in each ethnic group by country.

| **Country** | **Year** | **Ethnic group** | **%** | **95%CI** | | **Majority group (when applicable)** |
| --- | --- | --- | --- | --- | --- | --- |
| Afghanistan | 2015 | Other | 1·8 | 1·2 | 2·8 |  |
| Afghanistan | 2015 | Turkmen | 3·1 | 1·8 | 5 |  |
| Afghanistan | 2015 | Uzbek | 11·9 | 7·5 | 18·3 |  |
| Afghanistan | 2015 | Baloch | 0·6 | 0·3 | 1 |  |
| Afghanistan | 2015 | Hazara | 9·9 | 6·6 | 14·5 |  |
| Afghanistan | 2015 | Nuristani | 0·9 | 0·7 | 1·1 |  |
| Afghanistan | 2015 | Pashai | 1 | 0·6 | 1·6 |  |
| Afghanistan | 2015 | Pashtun | 40·3 | 36 | 44·8 |  |
| Afghanistan | 2015 | Tajik | 30·6 | 27·1 | 34·4 |  |
| Angola | 2015 | Other | 3·5 | 2·5 | 4·7 |  |
| Angola | 2015 | Chokwe/kioko | 5·1 | 4·1 | 6·2 |  |
| Angola | 2015 | Kikongo/ukongo | 4·8 | 3·8 | 6 |  |
| Angola | 2015 | Kwanhama | 2 | 1·5 | 2·7 |  |
| Angola | 2015 | Muhumbi | 1·6 | 0·7 | 3·3 |  |
| Angola | 2015 | Nganguela | 1·7 | 1·1 | 2·7 |  |
| Angola | 2015 | Nhaneca | 2 | 1 | 3·8 |  |
| Angola | 2015 | Português | 62·7 | 59·6 | 65·8 | yes |
| Angola | 2015 | Umbundu | 16·6 | 14·1 | 19·5 |  |
| Belize | 2015 | African descent | 27 | 22·4 | 32·3 |  |
| Belize | 2015 | Indigenous | 12·7 | 8·9 | 17·8 |  |
| Belize | 2015 | Reference | 60·3 | 54·4 | 65·9 | yes |
| Benin | 2017 | Other | 5·6 | 4 | 7·7 |  |
| Benin | 2017 | Adja and related | 12·4 | 11 | 14·1 |  |
| Benin | 2017 | Bariba and related | 12·3 | 9·6 | 15·6 |  |
| Benin | 2017 | Betamaribe and related | 6·6 | 5 | 8·6 |  |
| Benin | 2017 | Dendi and related | 5·4 | 3·7 | 7·9 |  |
| Benin | 2017 | Fon and related | 34·7 | 31·9 | 37·5 |  |
| Benin | 2017 | Peulh and related | 10·2 | 8 | 13 |  |
| Benin | 2017 | Yoa, Lokpa and related | 3·1 | 2·2 | 4·3 |  |
| Benin | 2017 | Yoruba | 9·7 | 8·1 | 11·6 |  |
| Bosnia and Herzegovina | 2011 | Bosnian | 66·2 | 60 | 71·8 | yes |
| Bosnia and Herzegovina | 2011 | Croat | 9 | 5·5 | 14·4 |  |
| Bosnia and Herzegovina | 2011 | Serb | 24·9 | 20·9 | 29·3 |  |
| Burkina Faso | 2010 | Bissa | 3·8 | 2·7 | 5·4 |  |
| Burkina Faso | 2010 | Bobo | 5·2 | 3·3 | 8 |  |
| Burkina Faso | 2010 | Dagara | 2·1 | 1·4 | 3·1 |  |
| Burkina Faso | 2010 | Fulfuldé / Peul | 9 | 7·2 | 11·3 |  |
| Burkina Faso | 2010 | Gourmatché | 8·9 | 7·2 | 10·9 |  |
| Burkina Faso | 2010 | Gourounsi | 4·5 | 3·2 | 6·4 |  |
| Burkina Faso | 2010 | Lobi | 2·4 | 1·6 | 3·7 |  |
| Burkina Faso | 2010 | Mossi | 49·9 | 46·4 | 53·4 |  |
| Burkina Faso | 2010 | Other | 7·1 | 5·6 | 9 |  |
| Burkina Faso | 2010 | Sénoufo | 4·7 | 3·1 | 7·1 |  |
| Burkina Faso | 2010 | Touareg / Bella | 2·4 | 1·5 | 3·7 |  |
| Central African Republic | 2018 | Banda | 19·5 | 16·5 | 22·9 |  |
| Central African Republic | 2018 | Gbaya | 32·9 | 28·7 | 37·4 |  |
| Central African Republic | 2018 | Haoussa | 3·2 | 2·1 | 4·9 |  |
| Central African Republic | 2018 | Mandja | 8·2 | 6 | 11·1 |  |
| Central African Republic | 2018 | Mboum | 8·5 | 5·1 | 13·8 |  |
| Central African Republic | 2018 | Ngbaka/Bantou | 6·8 | 4·9 | 9·3 |  |
| Central African Republic | 2018 | Other | 8 | 6·1 | 10·6 |  |
| Central African Republic | 2018 | Sara | 5·6 | 3·4 | 9 |  |
| Central African Republic | 2018 | Yakoma/Sango | 7·3 | 5·4 | 9·7 |  |
| Cameroon | 2018 | Other | 68 | 64 | 71·8 |  |
| Cameroon | 2018 | Bamileke | 7·7 | 6·2 | 9·5 |  |
| Cameroon | 2018 | Bamoun | 6·8 | 4·1 | 11·2 |  |
| Cameroon | 2018 | Boulou | 2·5 | 1·8 | 3·6 |  |
| Cameroon | 2018 | Eton | 3 | 1·9 | 4·8 |  |
| Cameroon | 2018 | Ewondo | 4 | 2·7 | 5·9 |  |
| Cameroon | 2018 | Foulbe | 7·9 | 5·9 | 10·4 |  |
| Chad | 2014 | Arab | 10·6 | 8·6 | 13·1 |  |
| Chad | 2014 | Other | 8·7 | 6·6 | 11·4 |  |
| Chad | 2014 | Sara | 31·8 | 28 | 35·8 |  |
| Chad | 2014 | Bidio/Migami/Kenga/Dangléat | 2·2 | 1·4 | 3·5 |  |
| Chad | 2014 | Boulala/Médégo/Kouka | 3·4 | 2·4 | 4·9 |  |
| Chad | 2014 | Dadajo/Kibet/Mouro | 2·5 | 1·6 | 4·1 |  |
| Chad | 2014 | Gorane | 4·8 | 3·8 | 6·1 |  |
| Chad | 2014 | Kanembou/Bornou/Boudouma | 7·7 | 6·4 | 9·3 |  |
| Chad | 2014 | Karo/Zimé/Pévé | 1·6 | 0·8 | 3 |  |
| Chad | 2014 | Marba/Lélé/Mesmé | 4·5 | 2·5 | 8 |  |
| Chad | 2014 | Massa/Mousseye/Mousgoume | 5·5 | 3·5 | 8·6 |  |
| Chad | 2014 | Moundang | 2·8 | 1·8 | 4·5 |  |
| Chad | 2014 | Ouadaï/Maba/Massalit/Mimi | 7·2 | 5·7 | 9 |  |
| Chad | 2014 | Peul/Foulbé/Bodoré | 2 | 1·3 | 3·1 |  |
| Chad | 2014 | Tama / Assongori / Mararit | 1·1 | 0·7 | 1·8 |  |
| Chad | 2014 | Toupouri/Kéra | 2·4 | 1·3 | 4·5 |  |
| Chad | 2014 | Zaghawa/Bideyat/Kobé | 1 | 0·6 | 1·6 |  |
| Colombia | 2010 | African descent | 12·4 | 11 | 14 |  |
| Colombia | 2010 | Indigenous | 6 | 5·1 | 7 |  |
| Colombia | 2010 | Reference | 81·6 | 79·9 | 83·2 | yes |
| Congo Brazzaville | 2014 | Kongo | 52·9 | 47·9 | 57·9 | yes |
| Congo Brazzaville | 2014 | Mbetis | 2·1 | 1·5 | 2·9 |  |
| Congo Brazzaville | 2014 | Mbosi | 12·4 | 9·6 | 15·9 |  |
| Congo Brazzaville | 2014 | Other | 15·1 | 12·4 | 18·2 |  |
| Congo Brazzaville | 2014 | Tekes | 11·9 | 9·7 | 14·4 |  |
| Congo Brazzaville | 2014 | Sangha-Likouala | 5·6 | 4·5 | 7 |  |
| Congo Democratic Republic | 2017 | Bantou | 93·2 | 90·5 | 95·2 | yes |
| Congo Democratic Republic | 2017 | Nilotique | 2·1 | 1·3 | 3·6 |  |
| Congo Democratic Republic | 2017 | Soudanais | 4·6 | 3·3 | 6·5 |  |
| Costa Rica | 2018 | African descent | 53·6 | 45·5 | 61·4 | yes |
| Costa Rica | 2018 | Reference | 46·4 | 38·6 | 54·5 |  |
| Cote dIvoire | 2016 | Akan | 24·8 | 21·4 | 28·5 |  |
| Cote dIvoire | 2016 | Gur | 17·2 | 14·8 | 20 |  |
| Cote dIvoire | 2016 | Krou | 8·7 | 6·5 | 11·5 |  |
| Cote dIvoire | 2016 | Mandé du Nord | 15·7 | 13·1 | 18·6 |  |
| Cote dIvoire | 2016 | Mandé du Sud | 7·8 | 5·8 | 10·4 |  |
| Cote dIvoire | 2016 | Other | 25·8 | 22·8 | 29·1 |  |
| Cuba | 2019 | African descent | 32·6 | 27·7 | 37·9 |  |
| Cuba | 2019 | Reference | 67·4 | 62·1 | 72·3 | yes |
| Dominican Republic | 2014 | African descent | 8·6 | 7·4 | 10 |  |
| Dominican Republic | 2014 | Reference | 91·4 | 90 | 92·6 | yes |
| Ethiopia | 2016 | Other | 23·7 | 20·2 | 27·7 |  |
| Ethiopia | 2016 | Affar | 1 | 0·7 | 1·5 |  |
| Ethiopia | 2016 | Amhara | 20·9 | 18·4 | 23·6 |  |
| Ethiopia | 2016 | Berta | 0·4 | 0·3 | 0·7 |  |
| Ethiopia | 2016 | Guragie | 1·4 | 0·8 | 2·4 |  |
| Ethiopia | 2016 | Nuwer | 0·1 | 0 | 0·1 |  |
| Ethiopia | 2016 | Oromo | 41·1 | 36·9 | 45·4 |  |
| Ethiopia | 2016 | Somalie | 3·6 | 2·8 | 4·6 |  |
| Ethiopia | 2016 | Tigrie | 7·7 | 6·4 | 9·2 |  |
| Gabon | 2012 | Other | 18·1 | 14·5 | 22·3 |  |
| Gabon | 2012 | Fang | 25·3 | 20·2 | 31·2 |  |
| Gabon | 2012 | Kota-Kele | 10·6 | 7·8 | 14·2 |  |
| Gabon | 2012 | Mbede-Teke | 9·6 | 6·8 | 13·4 |  |
| Gabon | 2012 | Nzabi-Duma | 16·6 | 12·8 | 21·2 |  |
| Gabon | 2012 | Shira-Punu/Vili | 19·9 | 16·1 | 24·2 |  |
| Gambia | 2018 | Fula | 21·9 | 18·4 | 26 |  |
| Gambia | 2018 | Jola | 10 | 7·4 | 13·4 |  |
| Gambia | 2018 | Mandinka | 29·7 | 25·7 | 34 |  |
| Gambia | 2018 | Other | 14·3 | 12 | 17 |  |
| Gambia | 2018 | Sarahule | 9·9 | 6·9 | 14 |  |
| Gambia | 2018 | Wollof | 14·1 | 11·3 | 17·5 |  |
| Ghana | 2017 | Akan | 47·5 | 42·6 | 52·5 |  |
| Ghana | 2017 | Ewe | 9·2 | 7·2 | 11·5 |  |
| Ghana | 2017 | Gruma | 4·6 | 3·2 | 6·7 |  |
| Ghana | 2017 | Grusi | 1·7 | 1·1 | 2·7 |  |
| Ghana | 2017 | Guan | 4·8 | 2·6 | 8·9 |  |
| Ghana | 2017 | Mole Dagbani | 17 | 13·1 | 21·8 |  |
| Ghana | 2017 | Other | 9·3 | 6·9 | 12·3 |  |
| Ghana | 2017 | Ga/Damgme | 5·8 | 4·4 | 7·6 |  |
| Guatemala | 2014 | Indigenous | 47 | 43·8 | 50·1 |  |
| Guatemala | 2014 | Reference | 53 | 49·9 | 56·2 | yes |
| Guinea | 2018 | Guerzé | 6·4 | 4·6 | 9 |  |
| Guinea | 2018 | Kissi | 5·7 | 3·7 | 8·8 |  |
| Guinea | 2018 | Malinké | 33·3 | 29·4 | 37·5 |  |
| Guinea | 2018 | Peulh | 34·9 | 31·1 | 38·9 |  |
| Guinea | 2018 | Soussou | 19·6 | 16·2 | 23·6 |  |
| Guinea Bissau | 2018 | Balanta | 21·3 | 17·6 | 25·4 |  |
| Guinea Bissau | 2018 | Beafada | 2·7 | 1·8 | 4·1 |  |
| Guinea Bissau | 2018 | Fula | 33·6 | 28·9 | 38·6 |  |
| Guinea Bissau | 2018 | Mandinga | 20·2 | 14·1 | 28·2 |  |
| Guinea Bissau | 2018 | Manjaco | 5·4 | 4 | 7·3 |  |
| Guinea Bissau | 2018 | Other | 9 | 6·9 | 11·7 |  |
| Guinea Bissau | 2018 | Papel | 7·8 | 6·1 | 9·9 |  |
| Guyana | 2014 | African descent | 30·8 | 25·7 | 36·4 |  |
| Guyana | 2014 | Indigenous | 15·1 | 11·6 | 19·5 |  |
| Guyana | 2014 | Reference | 54·1 | 48·4 | 59·7 | yes |
| Honduras | 2011 | African descent | 2·1 | 1·5 | 3 |  |
| Honduras | 2011 | Indigenous | 10 | 8·8 | 11·3 |  |
| Honduras | 2011 | Reference | 87·9 | 86·4 | 89·3 | yes |
| India | 2015 | Caste | 89 | 88·4 | 89·5 | yes |
| India | 2015 | No caste/Tribe | 3·8 | 3·4 | 4·2 |  |
| India | 2015 | Tribe | 7·2 | 6·9 | 7·6 |  |
| Iraq | 2018 | Arabic | 83·2 | 77·7 | 87·5 | yes |
| Iraq | 2018 | Kurdish | 16·8 | 12·5 | 22·3 |  |
| Jordan | 2017 | Other | 3·6 | 2·4 | 5·4 |  |
| Jordan | 2017 | Jordanian | 83·6 | 80·6 | 86·3 | yes |
| Jordan | 2017 | Syrian | 12·8 | 10·5 | 15·4 |  |
| Kazakhstan | 2015 | Kazakh | 73·3 | 67·8 | 78·1 | yes |
| Kazakhstan | 2015 | Other | 14·7 | 10·5 | 20·2 |  |
| Kazakhstan | 2015 | Russian | 12 | 9·9 | 14·4 |  |
| Kenya | 2014 | Other | 5·3 | 4·3 | 6·5 |  |
| Kenya | 2014 | Boran | 0·7 | 0·4 | 1·3 |  |
| Kenya | 2014 | Kalenjin | 13·3 | 11·8 | 15 |  |
| Kenya | 2014 | Kamba | 9·6 | 8·3 | 11·1 |  |
| Kenya | 2014 | Kikuya | 17·5 | 15·5 | 19·8 |  |
| Kenya | 2014 | Kisii | 5·3 | 4·3 | 6·5 |  |
| Kenya | 2014 | Luhya | 15·4 | 13·4 | 17·7 |  |
| Kenya | 2014 | Luo | 12·5 | 11·2 | 14 |  |
| Kenya | 2014 | Maasai | 3·4 | 2·5 | 4·6 |  |
| Kenya | 2014 | Meru | 4·2 | 3·4 | 5·2 |  |
| Kenya | 2014 | Mijikenda/Swahili | 5·9 | 5 | 7 |  |
| Kenya | 2014 | Pokomo | 0·4 | 0·2 | 0·7 |  |
| Kenya | 2014 | Samburu | 0·7 | 0·5 | 0·9 |  |
| Kenya | 2014 | Somali | 3·8 | 3·1 | 4·7 |  |
| Kenya | 2014 | Turkana | 1·9 | 1·4 | 2·6 |  |
| Kyrgyzstan | 2018 | Kyrgyz | 83·7 | 75·7 | 89·5 | yes |
| Kyrgyzstan | 2018 | Uzbek | 16·3 | 10·5 | 24·3 |  |
| Laos | 2017 | Chinese-Tibetan | 2·9 | 2 | 4 |  |
| Laos | 2017 | Hmong-Mien | 14·6 | 12·2 | 17·3 |  |
| Laos | 2017 | Lao-Tai | 57·1 | 53·4 | 60·7 | yes |
| Laos | 2017 | Mon-Khmer | 25·5 | 22·5 | 28·8 |  |
| Malawi | 2015 | Other | 3·9 | 3·2 | 4·7 |  |
| Malawi | 2015 | Chewa | 34·4 | 32·1 | 36·7 |  |
| Malawi | 2015 | Lomwe | 16·4 | 14·8 | 18·1 |  |
| Malawi | 2015 | Mang'Anja | 2·3 | 1·7 | 3·2 |  |
| Malawi | 2015 | Ngoni | 10·9 | 9·5 | 12·4 |  |
| Malawi | 2015 | Sena | 4·6 | 3·8 | 5·5 |  |
| Malawi | 2015 | Tonga | 1·9 | 1·5 | 2·6 |  |
| Malawi | 2015 | Tumbuka | 8·8 | 7·5 | 10·3 |  |
| Malawi | 2015 | Yao | 16·8 | 14·6 | 19·3 |  |
| Mali | 2018 | Other | 8 | 6·1 | 10·5 |  |
| Mali | 2018 | Bambara | 32 | 28 | 36·3 |  |
| Mali | 2018 | Dogon | 10·4 | 7·8 | 13·9 |  |
| Mali | 2018 | Malinke | 9·4 | 6·9 | 12·8 |  |
| Mali | 2018 | Peulh | 13·5 | 10·9 | 16·6 |  |
| Mali | 2018 | Sarakole/Soninke/Marka | 9·7 | 7·3 | 12·9 |  |
| Mali | 2018 | Sonraï | 5·7 | 4·6 | 7·1 |  |
| Mali | 2018 | Sénoufo/Minianka | 9·4 | 6·8 | 12·9 |  |
| Mali | 2018 | Touareg/Bella | 1·7 | 1 | 2·9 |  |
| Mauritania | 2015 | Arabe | 79·9 | 74·9 | 84·1 | yes |
| Mauritania | 2015 | Poular | 16·8 | 12·9 | 21·6 |  |
| Mauritania | 2015 | Soninké | 3·3 | 2 | 5·4 |  |
| Mexico | 2015 | Indigenous | 8·8 | 6·3 | 12 |  |
| Mexico | 2015 | Reference | 91·2 | 88 | 93·7 | yes |
| Mongolia | 2018 | Kazakh | 4·7 | 3·6 | 6·1 |  |
| Mongolia | 2018 | Khalkh | 78·8 | 75 | 82·2 | yes |
| Mongolia | 2018 | Other | 16·5 | 13·1 | 20·5 |  |
| Montenegro | 2013 | Montenegrin | 52·8 | 42·5 | 62·9 | yes |
| Montenegro | 2013 | Other | 25·9 | 17·6 | 36·3 |  |
| Montenegro | 2013 | Serbian | 21·3 | 14·6 | 30 |  |
| Mozambique | 2015 | Other | 29·6 | 24·4 | 35·3 |  |
| Mozambique | 2015 | Cindau | 5·5 | 3·1 | 9·6 |  |
| Mozambique | 2015 | Cinyanja | 5·7 | 3·1 | 10·5 |  |
| Mozambique | 2015 | Cisena | 6·6 | 4·3 | 10·1 |  |
| Mozambique | 2015 | Emakhuwa | 32 | 27·4 | 37 |  |
| Mozambique | 2015 | Português | 5·5 | 4 | 7·4 |  |
| Mozambique | 2015 | Xichangana | 10·4 | 8·7 | 12·4 |  |
| Mozambique | 2015 | Xitswa | 4·6 | 3·1 | 6·8 |  |
| Myanmar | 2015 | Other | 15·9 | 12·9 | 19·5 |  |
| Myanmar | 2015 | Myanmar | 84·1 | 80·5 | 87·1 | yes |
| Namibia | 2013 | Other | 6·1 | 4·3 | 8·5 |  |
| Namibia | 2013 | Afrikaans | 7 | 5 | 9·7 |  |
| Namibia | 2013 | Damara/Nama | 10·7 | 8·8 | 13·1 |  |
| Namibia | 2013 | Herero | 6·7 | 4·7 | 9·6 |  |
| Namibia | 2013 | Kwangali | 12·2 | 10·1 | 14·8 |  |
| Namibia | 2013 | Lozi | 5·6 | 4·6 | 6·9 |  |
| Namibia | 2013 | Oshiwambo | 51·6 | 47·8 | 55·4 | yes |
| Nepal | 2019 | Brahman - Hill | 11·3 | 9 | 14 |  |
| Nepal | 2019 | Chhetree | 14·7 | 12·4 | 17·3 |  |
| Nepal | 2019 | Kami | 4·5 | 3·4 | 5·9 |  |
| Nepal | 2019 | Magar | 6·8 | 5·1 | 9·1 |  |
| Nepal | 2019 | Musalman | 5·8 | 3·6 | 9·3 |  |
| Nepal | 2019 | Other | 44·5 | 40·3 | 48·9 |  |
| Nepal | 2019 | Tamang | 5·9 | 4·3 | 8·1 |  |
| Nepal | 2019 | Tharu | 6·5 | 4·5 | 9·1 |  |
| Niger | 2012 | Haoussa | 73·7 | 70·4 | 76·8 | yes |
| Niger | 2012 | Kanouri/Toubou | 1·9 | 1·5 | 2·5 |  |
| Niger | 2012 | Other | 3·4 | 2·5 | 4·6 |  |
| Niger | 2012 | Zarma | 21 | 18 | 24·3 |  |
| Nigeria | 2018 | Other | 22·5 | 20·7 | 24·6 |  |
| Nigeria | 2018 | Fulani | 8·1 | 6·8 | 9·6 |  |
| Nigeria | 2018 | Hausa | 34 | 31·6 | 36·4 |  |
| Nigeria | 2018 | Ibibio | 1·5 | 1·1 | 2 |  |
| Nigeria | 2018 | Igala | 0·7 | 0·5 | 1·1 |  |
| Nigeria | 2018 | Igbo | 13·8 | 12·6 | 15·2 |  |
| Nigeria | 2018 | Ijaw/Izon | 1·8 | 1·4 | 2·3 |  |
| Nigeria | 2018 | Kanuri/Beriberi | 2·5 | 1·8 | 3·5 |  |
| Nigeria | 2018 | Tiv | 2·6 | 1·9 | 3·4 |  |
| Nigeria | 2018 | Yoruba | 12·5 | 10·9 | 14·3 |  |
| North Macedonia | 2018 | Albanian | 37 | 26·3 | 49·3 |  |
| North Macedonia | 2018 | Macedonian | 63 | 50·7 | 73·7 | yes |
| Pakistan | 2017 | Other | 6·9 | 4·9 | 9·7 |  |
| Pakistan | 2017 | Baluchi | 2·8 | 1·8 | 4·3 |  |
| Pakistan | 2017 | Punjabi | 36·8 | 31·8 | 42·1 |  |
| Pakistan | 2017 | Pushto | 18·1 | 15 | 21·6 |  |
| Pakistan | 2017 | Sariaki | 15·9 | 11·5 | 21·4 |  |
| Pakistan | 2017 | Sindhi | 11·2 | 8·2 | 15 |  |
| Pakistan | 2017 | Urdu | 8·4 | 6·2 | 11·1 |  |
| Panama | 2013 | African descent | 14·4 | 11 | 18·5 |  |
| Panama | 2013 | Indigenous | 21 | 17·8 | 24·5 |  |
| Panama | 2013 | Reference | 64·7 | 59·4 | 69·6 | yes |
| Papua New Guinea | 2016 | English | 2·1 | 1·3 | 3·4 |  |
| Papua New Guinea | 2016 | Pidgin | 40·8 | 36·8 | 44·9 |  |
| Papua New Guinea | 2016 | Tok Ples | 57·1 | 52·9 | 61·1 | yes |
| Paraguay | 2016 | Indigenous | 41·7 | 37·1 | 46·4 |  |
| Paraguay | 2016 | Reference | 58·3 | 53·6 | 62·9 | yes |
| Peru | 2019 | Indigenous | 6·3 | 5·4 | 7·3 |  |
| Peru | 2019 | Reference | 93·7 | 92·7 | 94·6 | yes |
| Philippines | 2017 | Other | 21·5 | 19 | 24·2 |  |
| Philippines | 2017 | Bikolano | 6·5 | 5·3 | 8 |  |
| Philippines | 2017 | Cebuano | 19·2 | 16·9 | 21·7 |  |
| Philippines | 2017 | Ilokano | 6·8 | 5·2 | 8·8 |  |
| Philippines | 2017 | Ilonggo | 8·7 | 7·2 | 10·5 |  |
| Philippines | 2017 | Maranao | 2 | 1·4 | 2·9 |  |
| Philippines | 2017 | Tagalog | 27·7 | 24·4 | 31·2 |  |
| Philippines | 2017 | Visaya | 3·3 | 2·4 | 4·4 |  |
| Philippines | 2017 | Waray | 4·3 | 3·3 | 5·7 |  |
| Senegal | 2019 | Other | 10·5 | 7·5 | 14·6 |  |
| Senegal | 2019 | Poular | 29·8 | 25·7 | 34·2 |  |
| Senegal | 2019 | Wolof | 43·1 | 37·7 | 48·8 |  |
| Senegal | 2019 | Mandingue/ Socé | 5·2 | 3·7 | 7·5 |  |
| Senegal | 2019 | Serer | 11·4 | 8·3 | 15·4 |  |
| Sierra Leone | 2019 | Other | 12 | 9·8 | 14·6 |  |
| Sierra Leone | 2019 | Fullah | 2·7 | 2 | 3·7 |  |
| Sierra Leone | 2019 | Kono | 3·7 | 2·7 | 4·9 |  |
| Sierra Leone | 2019 | Korankoh | 4 | 2·6 | 6 |  |
| Sierra Leone | 2019 | Limba | 7·3 | 5·6 | 9·4 |  |
| Sierra Leone | 2019 | Mende | 34·8 | 31·7 | 38 |  |
| Sierra Leone | 2019 | Temne | 35·6 | 32·4 | 38·9 |  |
| South Africa | 2016 | Black/African | 91 | 88·2 | 93·2 | yes |
| South Africa | 2016 | Coloured | 9 | 6·8 | 11·8 |  |
| Suriname | 2018 | African descent | 35·7 | 30·4 | 41·4 |  |
| Suriname | 2018 | Reference | 64·3 | 58·6 | 69·6 | yes |
| Tajikistan | 2017 | Other | 16·3 | 11·8 | 22·2 |  |
| Tajikistan | 2017 | Russian | 83·7 | 77·8 | 88·2 | yes |
| Thailand | 2019 | Non-Thai | 8·6 | 6·7 | 11 |  |
| Thailand | 2019 | Thai | 91·4 | 89 | 93·3 | yes |
| Timor Leste | 2016 | Other | 6·1 | 4·6 | 8·1 |  |
| Timor Leste | 2016 | Tetum | 93·9 | 91·9 | 95·4 | yes |
| Togo | 2017 | Adja-Ewe | 36·8 | 31·6 | 42·2 |  |
| Togo | 2017 | Akposso/Akébou | 23·1 | 18·6 | 28·3 |  |
| Togo | 2017 | Kabye-Tem | 22·2 | 18·2 | 26·7 |  |
| Togo | 2017 | Other | 18 | 14 | 22·7 |  |
| Turkmenistan | 2015 | Turkmen | 91·3 | 86·6 | 94·5 | yes |
| Turkmenistan | 2015 | Uzbek | 8·7 | 5·5 | 13·4 |  |
| Uganda | 2016 | Other | 11·2 | 9·4 | 13·3 |  |
| Uganda | 2016 | Acholi | 4·3 | 3·5 | 5·2 |  |
| Uganda | 2016 | Alur | 3·2 | 2·1 | 4·7 |  |
| Uganda | 2016 | Bafumbira | 2·1 | 1·4 | 3·2 |  |
| Uganda | 2016 | Baganda | 15·2 | 13·1 | 17·5 |  |
| Uganda | 2016 | Bagisu | 4·9 | 4 | 6 |  |
| Uganda | 2016 | Bagwere | 1·9 | 1·3 | 2·9 |  |
| Uganda | 2016 | Bakiga | 5·9 | 4·7 | 7·3 |  |
| Uganda | 2016 | Bakonzo | 2·8 | 1·8 | 4·4 |  |
| Uganda | 2016 | Banyankore | 10·1 | 8·5 | 12·1 |  |
| Uganda | 2016 | Banyarwanda | 2·5 | 1·8 | 3·4 |  |
| Uganda | 2016 | Banyole | 2 | 1·3 | 3·1 |  |
| Uganda | 2016 | Banyoro | 2·9 | 2·1 | 4 |  |
| Uganda | 2016 | Basoga | 7·7 | 6·3 | 9·4 |  |
| Uganda | 2016 | Batoro | 3·4 | 2·4 | 4·9 |  |
| Uganda | 2016 | Iteso | 8·2 | 6·7 | 10·1 |  |
| Uganda | 2016 | Jopadhola | 2·3 | 1·5 | 3·6 |  |
| Uganda | 2016 | Karimojong | 1·2 | 0·7 | 2 |  |
| Uganda | 2016 | Lango | 5·2 | 4·3 | 6·3 |  |
| Uganda | 2016 | Lugbara | 3 | 2·2 | 4·1 |  |
| Vietnam | 2013 | Kinh | 85 | 81·5 | 88 | yes |
| Vietnam | 2013 | Non-Kinh | 15 | 12 | 18·5 |  |
| Zambia | 2018 | Other | 6·9 | 5·6 | 8·6 |  |
| Zambia | 2018 | Bemba | 36·8 | 33·6 | 40 |  |
| Zambia | 2018 | Kaonde | 2·8 | 1·9 | 4·2 |  |
| Zambia | 2018 | Lozi | 7·9 | 6·5 | 9·5 |  |
| Zambia | 2018 | Lunda | 2·2 | 1·4 | 3·5 |  |
| Zambia | 2018 | Luvale | 2·3 | 1·6 | 3·2 |  |
| Zambia | 2018 | Nyanja | 23·5 | 21 | 26·2 |  |
| Zambia | 2018 | Tonga | 17·6 | 14·5 | 21·1 |  |
| Zimbabwe | 2019 | Other | 4·9 | 2·8 | 8·4 |  |
| Zimbabwe | 2019 | Ndebele | 11·1 | 9·2 | 13·2 |  |
| Zimbabwe | 2019 | Shona | 84·1 | 80·9 | 86·8 | yes |

### Supplementary Table 6 – Median no-DPT prevalence by different grouping of countries.

| **Country groups** | **Number of countries** | **Median number of ethnic groups** | **Median no-DPT prevalence** | **Median high-low ethnic gap** | | **Pct countries with significant ethnic gaps*** |
| --- | --- | --- | --- | --- | --- | --- |
|  |  |  |  | **Difference (pp)** | **Ratio** |  |
| **GAVI eligibility status** |  |  |  |  |  |  |
| GAVI | 40 | 6·5 | 11·8 | 15·6 | 4·2 | 72·5 |
| Non-GAVI | 24 | 2 | 5·4 | 5·2 | 2·9 | 37·5 |
| **World region** |  |  |  |  |  |  |
| WCA | 20 | 6·5 | 14·1 | 17·9 | 3·4 | 70 |
| ESA | 10 | 8·5 | 6·4 | 12 | 9·1 | 60 |
| MENA | 2 | 2·5 | 10·4 | 10·5 | 2·6 | 50 |
| EECA | 7 | 2 | 4·4 | 1·6 | 1·8 | 28·6 |
| SA | 4 | 7·5 | 12·1 | 22·1 | 5·8 | 100 |
| EAP | 8 | 2·5 | 14·3 | 14·3 | 3·8 | 75 |
| LAC | 13 | 2 | 4·7 | 3 | 2·8 | 38·5 |
| **Country income group** |  |  |  |  |  |  |
| Low | 20 | 8 | 10·3 | 20·5 | 6·3 | 80 |
| Lower middle | 23 | 3 | 13·1 | 10·6 | 3·3 | 69·6 |
| Upper middle | 21 | 2 | 5·2 | 3·7 | 2 | 28·6 |
| **Number of ethnic groups** |  |  |  |  |  |  |
| <4 | 33 | 2 | 5·6 | 5·5 | 2·2 | 45·5 |
| 4 to 8 | 18 | 6·5 | 10·3 | 14·6 | 3·5 | 61·1 |
| >8 | 13 | 9 | 17·9 | 43·5 | 8·5 | 92·3 |
| **No-DPT prevalence** |  |  |  |  |  |  |
| <5% | 20 | 3 | 3 | 3·7 | 4·3 | 45 |
| 5-9% | 17 | 3 | 7·3 | 8·9 | 2·8 | 35·3 |
| >=10% | 27 | 6 | 17·9 | 23·9 | 3·5 | 85·2 |
| **All countries** | 64 | 3 | 7·8 | 9·6 | 3·3 | 59·4 |

Legend: WCA: West and Central Africa. ESA: Eastern and South Africa. MENA: Middle East and North Africa. ECA: Europe and Central Asia. SA: South Asia. EAP: East Asia and The Pacific. LAC: Latin America and Caribbean. No-DPT: children who did not received any doses of the diphtheria-tetanus-pertussis-containing vaccine. *p value < 0·05.
